# Trajectories of metabolic and inflammatory biomarkers ten years before cancer diagnosis and the risk of subsequent severe infections: a Swedish population-based cohort study

**DOI:** 10.1101/2025.09.23.25336508

**Authors:** Dang Wei, Donghang Zhang, Quan Wang, Qiaoxue Liu, Niklas Hammar, Kejia Hu, Maria Feychting, Kewei Jiang, Qianwei Liu, Fang Fang

**Affiliations:** Institute of Environmental Medicine, Karolinska Institutet, Stockholm, Sweden; Department of Social and Behavioral Sciences, Harvard T.H. Chan School of Public Health, Boston, MA, USA; Department of Anesthesiology, West China Hospital, Sichuan University, Chengdu, China; Laboratory of Anesthesia and Critical Care Medicine, National-Local Joint Engineering Research Centre of Translational Medicine of Anesthesiology, West China Hospital, Sichuan University, Chengdu, China; Ambulatory Surgery Center, Xijing Hospital, the Fourth Military Medical University, Xi’an, China; Department of Hematology, Nanfang Hospital, Southern Medical University, Guangzhou, China; Clinical Medical Research Center of Hematology Diseases of Guangdong Province, Guangzhou, China; Department of Gastrointestinal Surgery, Laboratory of Surgical Oncology, Beijing Key Laboratory of Colorectal Cancer Diagnosis and Treatment Research, Peking University People’s Hospital, Beijing, China

**Author notes:** Corresponding to: Qianwei Liu, Department of Hematology, Nanfang Hospital, Southern Medical University, No.1838, North Guangzhou Avenue, Guangzhou, 510515, China. DW, DHZ, QW, and QXL contributed equally to the work.

**Keywords:** Cancer, infection, longitudinal trajectories, metabolic biomarkers, inflammatory biomarkers

## Abstract

**Background:** This study investigated longitudinal trajectories of circulating metabolic and inflammatory biomarkers before cancer diagnosis and assess their associations with the risk of subsequent severe infections.

**Methods:** We conducted a cohort study including 10,837 patients with a first diagnosis of cancer during 1985-2005 from a Swedish cohort. We studied 18 biomarkers and used latent class growth modeling for identifying biomarker trajectories within 10 years before cancer diagnosis and flexible parametric models for assessing their associations with severe infections.

**Results:** Compared to patients with persistently low levels, those with persistently high levels of triglycerides [HR (95%CI): 1.27 (1.14–1.41)] and haptoglobin [1.38 (1.23–1.55)] or increasing CRP levels within five years before cancer diagnosis [1.24 (10.6-1.46)] had a higher risk of hospitalization for infectious diseases. However, persistently low HDL levels were associated with an increased risk. Further, dose-response associations were observed for uric acid and albumin, which persistently higher uric acid levels and lower albumin levels were associated with a higher risk of hospitalization for infectious diseases. Similar results were observed for sepsis.

**Conclusion:** Temporal trajectories in metabolic and inflammatory biomarkers during the 10 years before cancer diagnosis were linked to an altered risk of subsequent severe infections.

## Introduction

Cancer represents a major global public health challenge, with an estimated 20 million new cases diagnosed worldwide in 2022, accounting for approximately one in six deaths.(1) Severe infections, including sepsis, are common and serious complications in cancer patients. These infections can interrupt oncologic treatment, accelerate clinical deterioration, and substantially increase mortality.(2-4) Evidence from autopsy studies suggests that infections may account for up to 60% of deaths among patients with hematological malignancies,(2, 5) underscoring their critical contribution to cancer-related mortality.

Identifying patients at heightened risk of severe infections is therefore essential for improving cancer survivorship. The risk of sepsis—one of the most severe manifestations of infection— has been shown to peak in the period immediately following a cancer diagnosis.(6) This temporal pattern highlights the clinical importance of early risk stratification at the time of diagnosis, ideally using indicators applicable across cancer types. Several pre-diagnostic factors have been associated with an increased risk of post-diagnostic infections, including immune dysfunction, pre-existing comorbidities such as diabetes and psychiatric disorders, poor nutritional status, and prior antimicrobial use.(2, 3, 6) Collectively, these findings point to a central role of immune and metabolic dysregulation in susceptibility to severe infections among patients with cancer.

Despite this, knowledge remains limited regarding whether circulating metabolic and inflammatory biomarkers measured before cancer diagnosis are associated with the subsequent risk of severe infections in a pan-cancer context. Previous studies have largely focused on specific cancer types.(7, 8) For instance, declining levels of total cholesterol (TC) and low-density lipoprotein (LDL), alongside increasing glucose levels, have been observed in the years preceding cancer diagnosis compared with cancer-free individuals.(7, 8) Whether such patterns are consistent across different cancer types, and whether they carry prognostic information for infection risk after diagnosis, remains unclear.

Importantly, metabolic and inflammatory biomarkers are dynamic and may change substantially over time. Reliance on single pre-diagnostic measurements may therefore inadequately capture underlying biological processes and lead to biased estimates of disease risk.(9, 10) Longitudinal trajectories of biomarkers may provide more informative insights into chronic metabolic and inflammatory alterations preceding cancer onset.(9, 10) However, to the best of our knowledge, the extent to which pre-diagnostic temporal trajectories of metabolic and inflammatory biomarkers are associated with the risk of severe infections following a cancer diagnosis has rarely been examined.

To address these gaps, we used the Swedish AMORIS (Apolipoprotein-related MOrtality RISk) cohort to investigate the longitudinal trajectories of 18 metabolic and inflammatory biomarkers in blood during the 10 years before a cancer diagnosis, and to evaluate their associations with the risk of severe infections following the cancer diagnosis.

## Materials and methods

### Study design and population

The Swedish AMORIS cohort includes 812,073 individuals who underwent blood or urine laboratory tests for outpatient visits or regular health check-ups in the occupational setting during 1985-1996 in Stockholm.(11) A total of 634,981 individuals had data on blood tests during the inclusion period. Through the unique personal identification number assigned to each Swedish resident, the AMORIS cohort has been linked to several national population and health registers, including the Swedish Cancer Register, the Swedish Patient Register, the Causes of Death Register, and the Total Population Register. Cancer diagnoses were identified in the Cancer Register through the end of 2020, using the Swedish adaptation of the seventh revision of the International Classification of Diseases (ICD), which has been in use since the register was established (Table S1). As we focused on biomarker trajectories within 10 years before a cancer diagnosis, we conducted a cohort study including patients who were diagnosed with their first cancer during 1985-2005 in the AMORIS cohort. To evaluate quadratic changes in temporal patterns of biomarker trajectories, we restricted the analysis to cancer patients who had undergone at least three health examinations with blood biomarker measurements prior to diagnosis, yielding 10,837 eligible patients (Figure 1). No substantial differences were found between patients included in this analysis and cancer patients diagnosed during 1985-2005 with fewer than three blood tests prior to cancer diagnosis (Table S2). We then followed all eligible patients from the date of cancer diagnosis to a first event of severe infection (identified through the Patient Register), emigration (identified through the Total Population Register), death (identified through the Causes of Death Register), or December 31, 2020, whichever came first.

**Figure 1.**
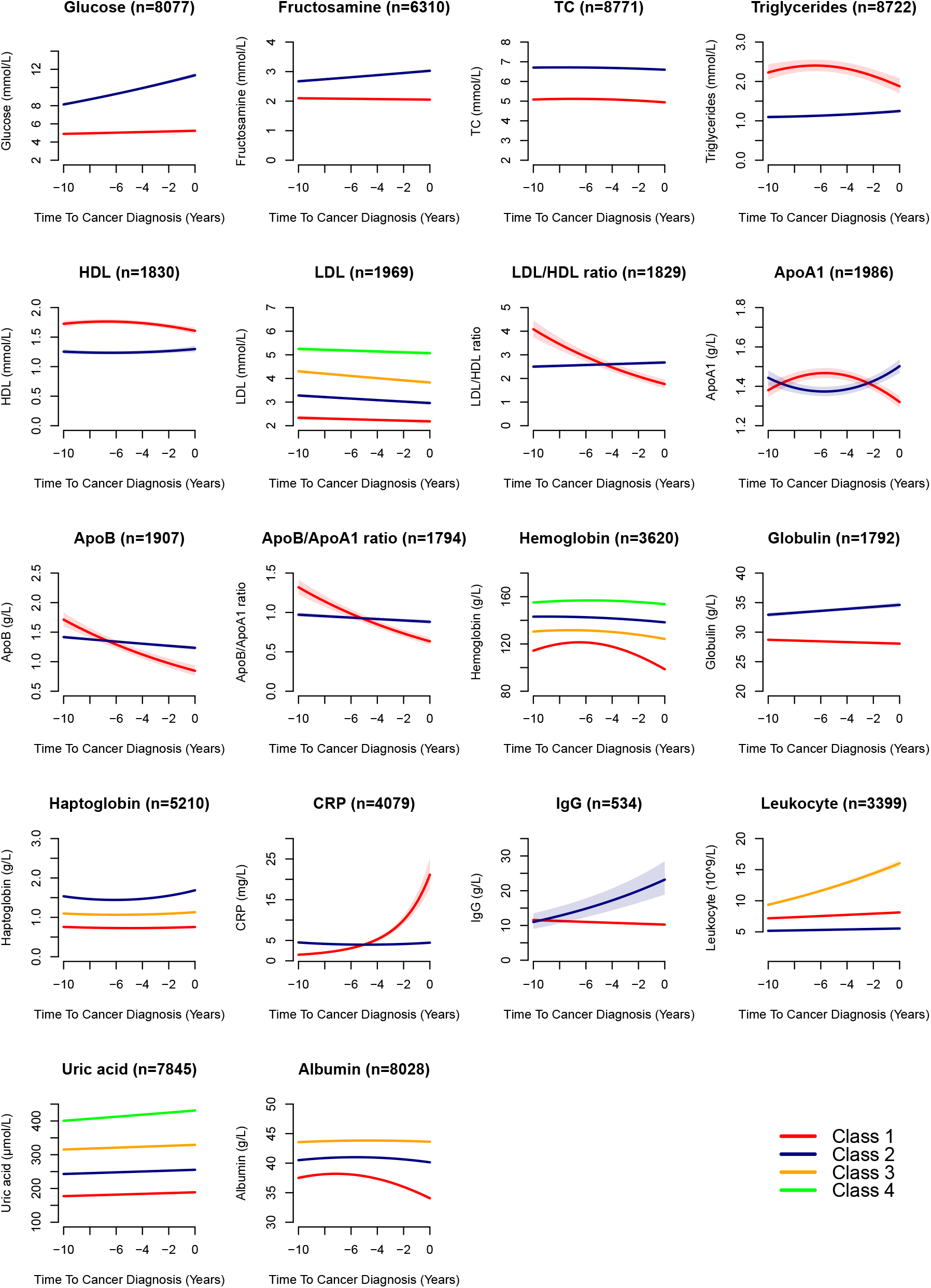
Flowchart of study design based on the Swedish AMORIS cohort.

### Metabolic and inflammatory biomarkers

We focused on 18 metabolic and inflammatory blood biomarkers which were commonly measured in the AMORIS cohort (i.e., available among at least 20% of the cohort population), including glucose, fructosamine, TC, high-density lipoprotein (HDL), LDL, LDL/HDL ratio, triglycerides, apolipoprotein A1 (ApoA1), apolipoprotein B (ApoB), ApoB/ApoA1 ratio, albumin, uric acid, haptoglobin, leukocyte, immunoglobulin G (IgG), C-reactive protein (CRP), hemoglobin, and globulin. LDL concentrations were mainly calculated using the Jungner formula, while HDL concentrations were derived from TC, triglycerides, and ApoA-I levels for most individuals and measured directly in blood for a few others.(12)

### Severe infections

We defined the primary outcome as a hospitalization for infectious diseases and the secondary outcome as a diagnosis of sepsis through the Swedish Patient Register. As milder infections will generally not lead to hospitalization and may only be recorded as secondary diagnoses in inpatient care, we identified hospitalizations for infectious diseases using the primary diagnosis of an inpatient care episode only. In contrast, sepsis was identified using both primary and secondary diagnoses from both inpatient and outpatient care. Table S1 shows the Swedish versions of the ICD codes used to ascertain these outcomes.

### Covariates

We obtained information on sex and birth year from the AMORIS cohort laboratory data, as well as data on country of birth, education, income, and employment status from the Swedish Population and Housing Census (FOB) in 1985 and the Longitudinal Integration Database for Health Insurance and Labour Market Studies (LISA) with annually updated information from 1990 onward. As education, income, and employment status were considered as potential confounders for the associations between biomarker trajectories within 10 years prior to cancer diagnosis and risk of severe infections after cancer diagnosis, we defined these covariates using available data closest in time to the first blood sampling during this time period. Individuals with missing values in these covariates were classified into an unknown category. We considered severe infections (both hospitalization for infectious diseases and a diagnosis of sepsis) during the 10 years before cancer diagnosis as well as history of diabetes and psychiatric disorders any time before cancer diagnosis as additional covariates.

### Statistical analysis

We first used latent class growth modeling (LCGM) to identify latent classes of trajectories of the 18 selected biomarkers during the 10 years before cancer diagnosis. We conducted three crude models with two polynomial functions of time (linear and quadratic) for biomarker trajectories: fixed effect only, fixed effect with random intercept, and fixed effect with both random intercept and slope. We analyzed one to five classes in each polynomial model. A log-transformation was used for all biomarkers in the LCGM analysis. The best-fit model was determined if the model had the lowest BIC with P<0.05 for all polynomial terms and a group membership probability of ≥5%. At the end, quadratic models were selected for ApoA1, triglycerides, HDL, hemoglobin, albumin, haptoglobin, and CRP, while linear models were selected for the other biomarkers (Table S3).

We then employed flexible parametric models to visualize hazards and hazard ratios (HRs) of severe infections in relation to pre-diagnostic biomarker trajectories by time since cancer diagnosis. Flexible parametric models allow for evaluating time-varying effect of exposure. As HRs were largely constant during the follow-up for most biomarker trajectories (Figure S1 for hospitalization for infectious diseases), we reported average HRs and 95% confidence intervals (CIs) for the sake of simplicity. Since the associations for glucose, fructosamine, TC, and leukocyte varied by time, we reported corresponding average HRs and 95% CIs after controlling for time-varying effects in the models. The classes with the largest number of cancer patients were defined as reference, whereas time since cancer diagnosis was used as the underlying time scale in the analyses. We conducted a multivariable model with the adjustment for sex, age and calendar year at cancer diagnosis, and country of birth, education, income, and employment status at the time of first blood sampling. To minimize potential confounding by comorbidities, we performed a sensitivity analysis by additionally adjusting for severe infections within the 10 years before cancer diagnosis, as well as any history of diabetes or psychiatric disorders before cancer diagnosis. To assess the robustness of the results for the ascertainment of hospitalization for infectious diseases via primary diagnosis only, we conducted a sensitivity analysis to consider both primary and secondary diagnoses from inpatient care when defining hospitalization for infectious diseases.

To evaluate if the associations between biomarker trajectories and severe infections differed by sex, age at cancer diagnosis, and history of severe infections during the 10 years before cancer diagnosis, we performed stratified analyses by these factors. Finally, to examine whether the results were specific to cancer types, we investigated the associations among patients with the three most common types of cancer in this cohort, namely cancer in breast or reproductive system, cancer in the digestive system, and hematological malignancies, separately.

The study was approved by the Swedish Ethical Review Authority (2018/2401-31, with subsequent amendments). Analyses were performed in SAS 9.4 and R 4.3.1.

## Results

Among the 10,837 cancer patients included in the study [mean age at diagnosis (standard deviation): 65.9 (11.7) years], 5476 (50.5%) were men, 4185 (38.6%) were diagnosed during 1985-1995, and 9385 (86.6%) were born in Sweden (Table 1). During the follow-up, 3752 (34.6%) of these patients had at least one hospitalization for infectious diseases whereas 1709 (15.8%) had a diagnosis of sepsis. Compared to patients without any hospitalization for infectious diseases by the end of follow-up, those with such experience were more likely to be employed, have higher income, and have had a severe infection during the 10 years before cancer diagnosis.

**Table 1.** Characteristics of the cancer patients included in the present study according to hospitalization for infectious disease during follow-up.

| Characteristics | Hospitalization<br>for infectious<br>disease<br>(N=3752) | No hospitalization<br>for infectious<br>disease<br>(N=7085) | Overall<br>(N=10,837) |
| --- | --- | --- | --- |
| Sex: male, N (%) | 1920 (51.2) | 3556 (50.2) | 5476 (50.5) |
| Age at diagnosis, mean (SD) | 65.7 (11.3) | 66.0 (11.9) | 65.9 (11.7) |
| Calendar year at diagnosis, N (%) |  |  |  |
| 1985-1995 | 1347 (35.9) | 2838 (40.1) | 4185 (38.6) |
| 1996-2005 | 2405 (64.1) | 4247 (59.9) | 6652 (61.4) |
| Country of birth, N (%) |  |  |  |
| Sweden | 3256 (86.8) | 6129 (86.5) | 9385 (86.6) |
| Elsewhere | 496 (13.2) | 956 (13.5) | 1452 (13.4) |
| Education at the first blood sampling, N (%) |  |  |  |
| Primary | 859 (22.9) | 1664 (23.5) | 2523 (23.3) |
| Secondary | 1819 (48.5) | 3241 (45.7) | 5060 (46.7) |
| Tertiary | 919 (24.5) | 1537 (21.7) | 2456 (22.7) |
| Unknown | 155 (4.1) | 643 (9.1) | 798 (7.4) |
| Employment status at the first blood sampling,<br>N (%) |  |  |  |
| Non-gainfully employed or unemployed | 926 (24.7) | 1987 (28.0) | 2913 (26.9) |
| Gainfully employed | 2811 (74.9) | 5056 (71.4) | 7867 (72.6) |
| Unknown | 15 (0.4) | 42 (0.6) | 57 (0.5) |
| Income at the first blood sampling, N (%) |  |  |  |
| Below 1st tertile | 1235 (32.9) | 2533 (35.8) | 3768 (34.8) |
| 1st-2nd tertile | 1093 (29.1) | 2103 (29.7) | 3196 (29.5) |
| Above 2nd tertile | 1417 (37.8) | 2432 (34.3) | 3849 (35.5) |
| Unknown | 7 (0.2) | 17 (0.2) | 24 (0.2) |
| Severe infections during the 10 years before<br>cancer diagnosis: yes, N (%)* | 540 (14.4) | 840 (11.9) | 1380 (12.7) |
| Psychiatric disorders before cancer diagnosis:<br>yes, N (%) | 304 (8.1) | 603 (8.5) | 907 (8.4) |
| Diabetes before cancer diagnosis: yes, N (%) | 248 (6.6) | 476 (6.7) | 724 (6.7) |

SD = standard deviation
\*Severe infections included hospitalization for infectious diseases and any diagnosis of sepsis at a hospital visit.

### Biomarker trajectories during the 10 years before cancer diagnosis

We identified two latent classes for most metabolic biomarkers, except for LDL, which exhibited four parallel trajectories (Figure 2). The trajectories of glucose, fructosamine, TC, triglycerides, and HDL showed consistently high or low levels during the 10 years before cancer diagnosis. The trajectories of LDL/HDL ratio, ApoB, and ApoB/ApoA1 ratio showed different patterns, with one trajectory showing stable levels whereas the other showing initially high levels followed by a constant decline over time. The two trajectories of ApoA1 were close to each other throughout the 10-year period.

**Figure 2.**
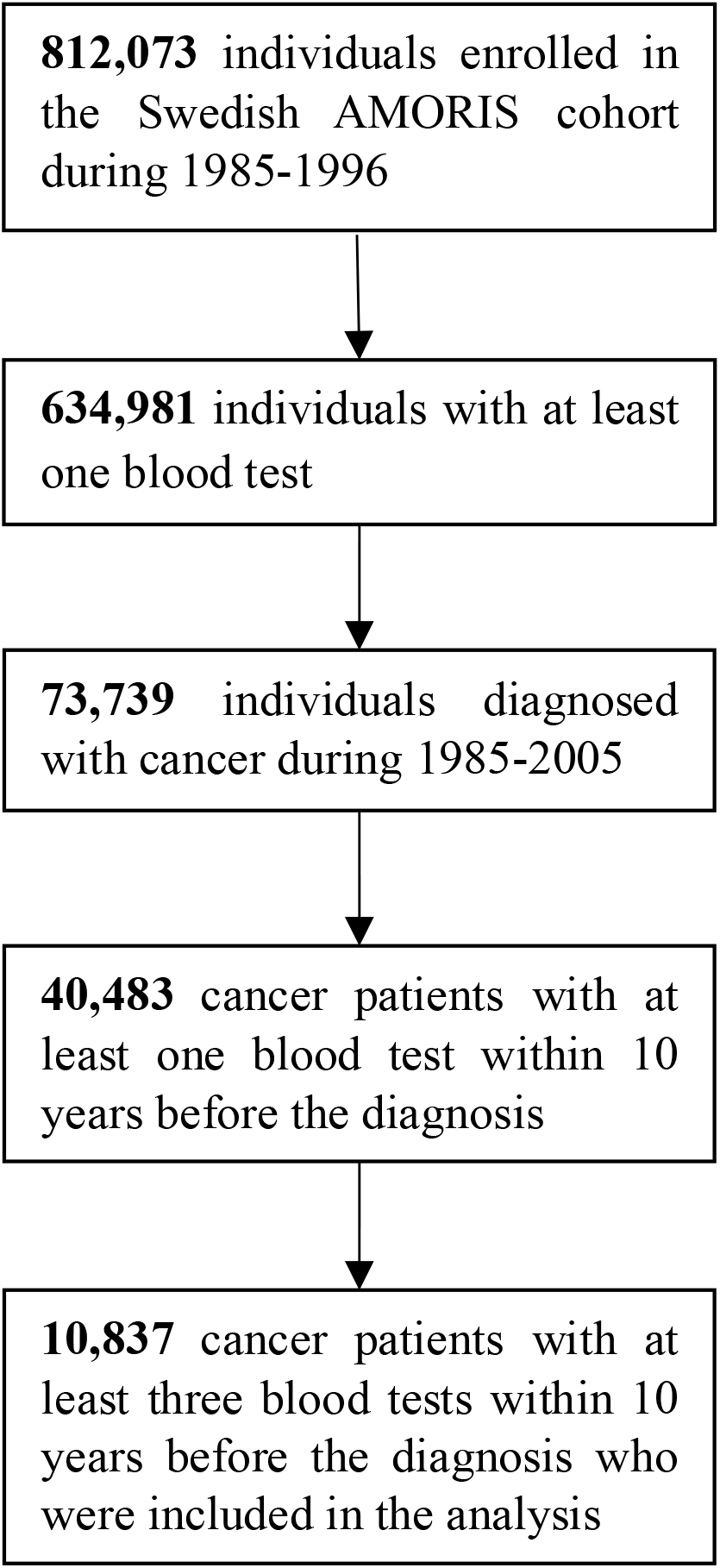
Latent classes of metabolic and inflammatory biomarker trajectories within 10 years before cancer diagnosis. TC=total cholesterol; HDL=high-density lipoprotein; LDL=low-density lipoprotein; ApoA1=apolipoprotein A1; ApoB=apolipoprotein B; CRP=C-reactive protein; IgG=immunoglobulin G Glucose (Class 1: stable at low levels; Class 2: started at high levels and increased); Fructosamine (Class 1: stable at low levels; Class 2: started at high levels and slightly increased); TC (Class 1: stable at low levels; Class 2: stable at high levels); Triglycerides (Class 1: started at high levels, remained stable, then decreased; Class 2: stable at low levels); HDL (Class 1: started at high levels, remained stable, then slightly decreased; Class 2: stable at low levels); LDL (Class 1: stable at low levels; Class 2: stable at low-middle levels; Class 3: stable at high-middle levels; Class 4: stable at high levels); LDL/HDL ratio (Class 1: started at high levels and decreased; Class 2: stable at low levels); ApoA1 (Class 1: slight increase followed by a decrease; Class 2: slight decrease followed by an increase); ApoB (Class 1: started at high levels and decreased; Class 2: stable at low levels); ApoB/ApoA1 ratio (Class 1: started at high levels and decreased; Class 2: stable at low levels); Hemoglobin (Class 1: started at low levels, remained stable, then decreased; Class 2: stable at low-middle levels; Class 3: stable at high-middle levels; Class 4: stable at high levels); Globulin (Class 1: stable at low levels; Class 2: started at high levels and slightly increased); Haptoglobin (Class 1: stable at low levels; Class 2: stable at high levels; Class 3: stable at intermediate levels); CRP (Class 1: started low levels, remained stable, then increased rapidly; Class 2: started at relatively higher levels but remained stable); IgG (Class 1: stable at low levels; Class 2: started at low levels and increased rapidly); Leukocyte (Class 1: stable at intermediate levels; Class 2: stable at low levels; Class 3: started at high levels and increased); Uric acid (Class 1: stable at low levels; Class 2: stable at low-middle levels; Class 3: stable at high-middle levels; Class 4: stable at high levels); Albumin (Class 1: started at low levels, remained stable, then decreased; Class 2: stable at intermediate levels; Class 3: stable at high levels).

Two distinct trajectories were identified for globulin, CRP, and IgG, whereas three trajectories were identified for haptoglobin, leukocyte, and albumin, and four trajectories were found for hemoglobin and uric acid. Globulin exhibited two trajectories with consistently high or low levels over time, whereas CRP and IgG showed one trajectory with stable levels and another with increasing levels over time. Trajectories of hemoglobin, haptoglobin, and uric acid displayed were largely parallel over time. Leukocytes showed one trajectory of increasing levels and two trajectories of stable levels, whereas albumin showed two trajectories with relatively high and stable levels but one trajectory that started at low levels with further decline over time.

### Associations between biomarker trajectories and severe infections after cancer diagnosis

The hazard of severe infections defined as either hospitalization for infectious diseases (Figure 3) or a diagnosis of sepsis (Figure S2) was highest shortly after cancer diagnosis, followed by a marked decrease during the first five years and a continuous increase during the 5-35 years after cancer diagnosis.

**Figure 3.**
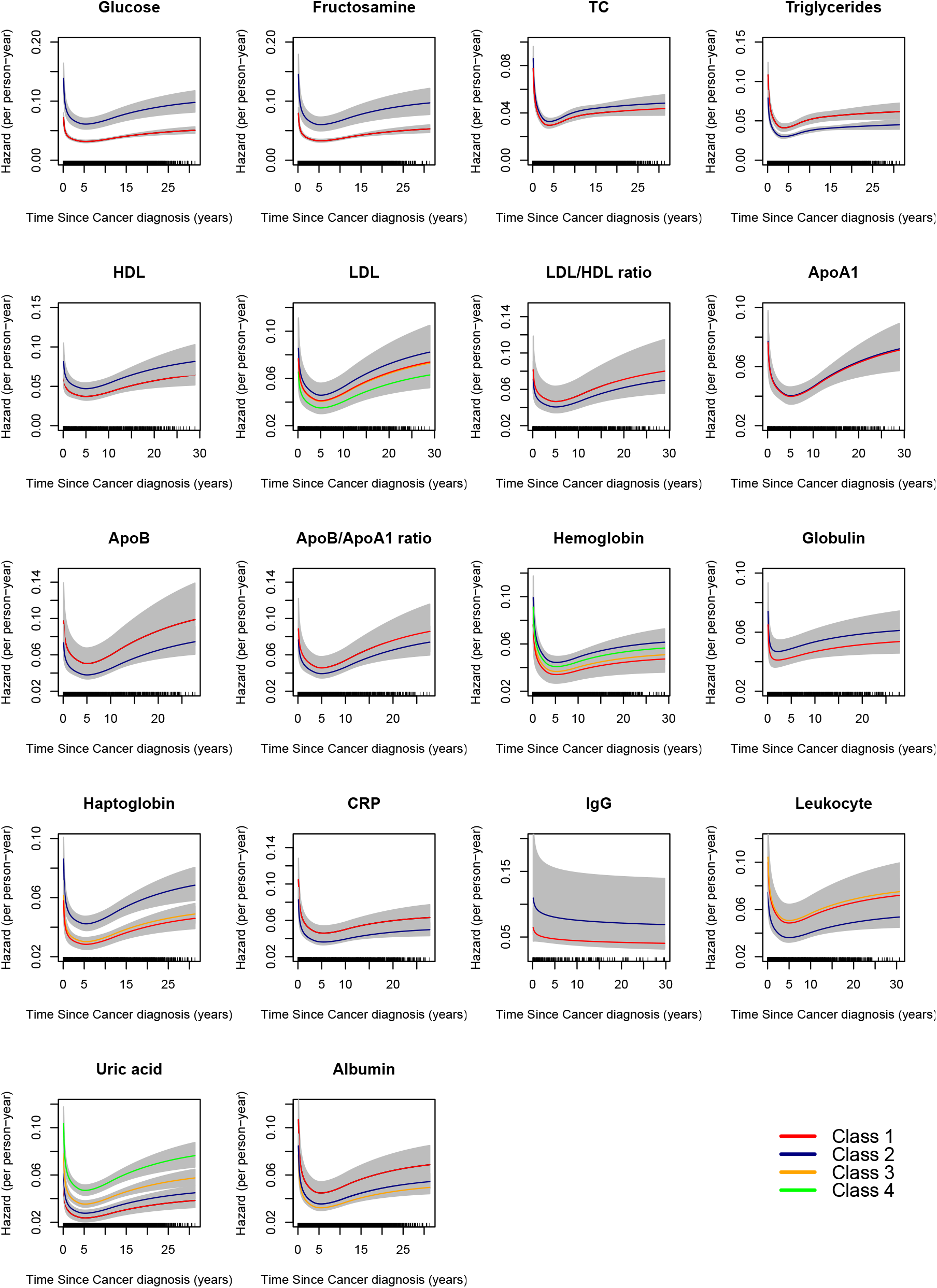
Hazards of hospitalization for infectious diseases after a cancer diagnosis in relation to biomarker trajectories. TC=total cholesterol; HDL=high-density lipoprotein; LDL=low-density lipoprotein; ApoA1=apolipoprotein A1; ApoB=apolipoprotein B; CRP=C-reactive protein; IgG=immunoglobulin G

We observed that cancer patients with persistently high triglycerides levels during the 10 years before diagnosis had a 27% higher risk of hospitalization for infectious diseases [HR (95% CI): 1.27 (1.14-1.41)] and a 26% higher risk of sepsis diagnosis [1.26 (1.08-1.46)], compared to those with persistently low levels during the 10 years before cancer diagnosis (Figure 4). In contrast, cancer patients with persistently low HDL levels had a 26% higher risk of hospitalization for infectious diseases [1.26 (1.07-1.49)]. No association was found for the trajectories of other metabolic biomarkers (Table S4).

**Figure 4.**
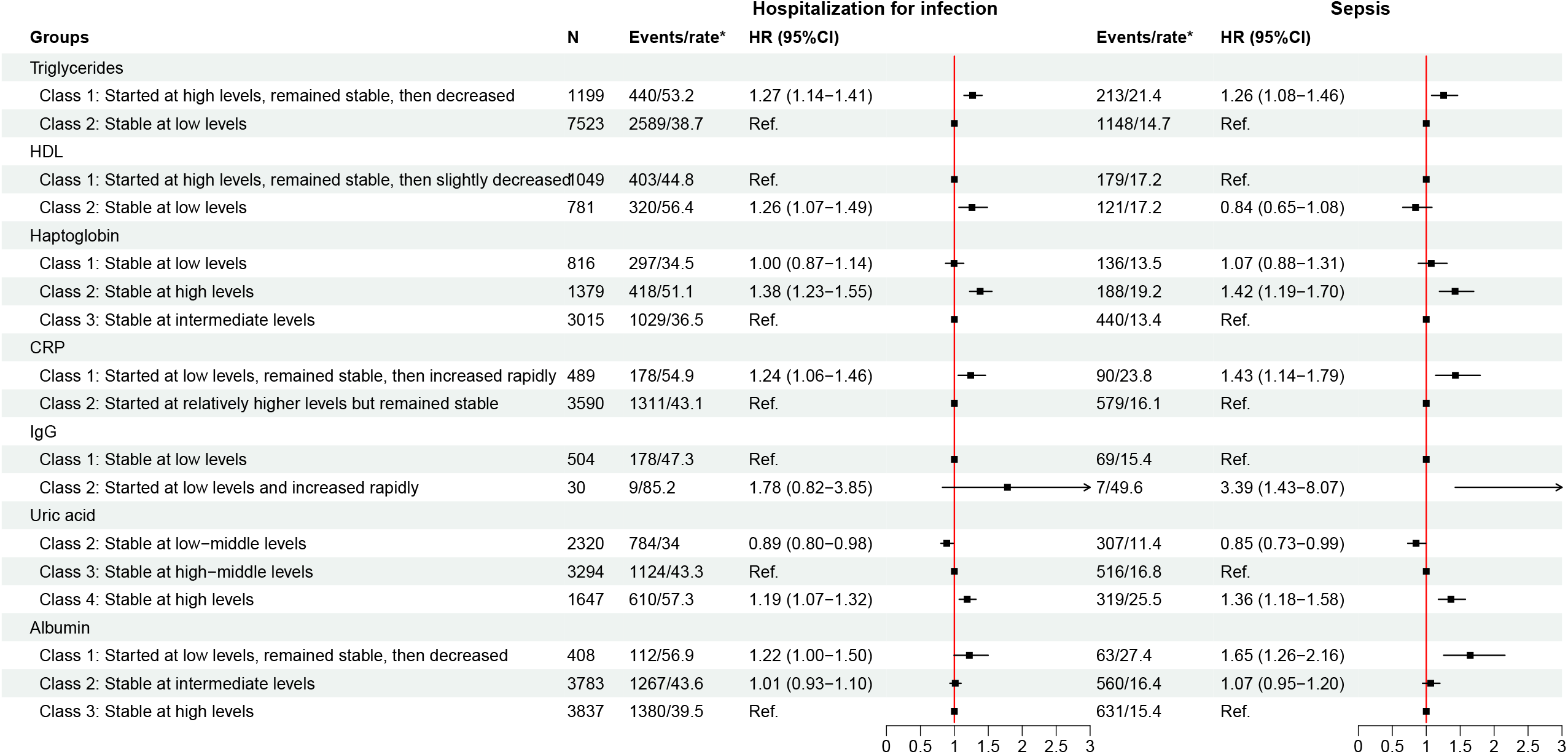
Associations of biomarker trajectories with hospitalization for infectious disease and sepsis. TC=total cholesterol; HDL=high-density lipoprotein; LDL=low-density lipoprotein; ApoA1=apolipoprotein A1; ApoB=apolipoprotein B; CRP=C-reactive protein; IgG=immunoglobulin G Adjusted for sex, calendar year of cancer diagnosis, age at cancer diagnosis, country of birth, education, income, and employment status at the first blood sampling. *rate: per 1000 person-years.

Cancer patients with persistently high levels of haptoglobin during the 10 years before cancer diagnosis had about 40% higher risk of severe infections than those with persistently low levels (Figure 4). Patients whose CRP or IgG levels were initially low but rapidly increased during the five or 10 years prior to cancer diagnosis had an elevated risk of severe infections, compared to those with persistently low levels. Dose-response associations were observed for trajectories of uric acid and albumin. Specifically, patients with persistently higher levels of uric acid preceding cancer diagnosis exhibited a higher risk of severe infections, whereas those with persistently lower albumin levels showed a higher risk (Figure 4). We did not find any association for other inflammatory biomarkers (Table S4).

We found similar results after additionally adjusting for history of severe infections during the 10 years before cancer diagnosis and psychiatric disorders and diabetes any time before cancer diagnosis (Table S5), or when considering both primary and secondary diagnoses for the ascertainment of hospitalization for infectious diseases (Table S6).

Compared to men, women had a greater risk increase in hospitalization for infectious diseases in relation to persistently high trajectories of triglycerides, haptoglobin, and uric acid as well as the persistently low trajectory of HDL (Table S7). In contrast, the risk decrease in relation to trajectories of persistently low and low-middle uric acid levels was larger in women than men. The risk decrease in relation to persistently high-middle hemoglobin levels and persistently low-middle uric acid levels was larger in patients diagnosed with cancer at age 60 or younger, compared to patients diagnosed later in life (Table S8). No substantial difference was observed for the associations according to history of severe infections at cancer diagnosis (Table S9) and cancer types i.e., breast and reproductive system cancers, digestive system cancers, and hematological malignancies (Table S10).

## Discussion

In this large Swedish cohort study, we identified several distinct trajectories for 18 metabolic and inflammatory blood biomarkers during the 10 years before a cancer diagnosis. Patients with consistently high levels of triglycerides during the 10 years prior to cancer diagnosis had a higher risk of severe infections, compared to those with persistently low levels. Conversely, patients with persistently low HDL levels exhibited a slightly higher risk of severe infections than those with persistently high levels. Increasing CRP and IgG levels, particularly during the five years before diagnosis, were associated with a higher risk of severe infections. Furthermore, a higher risk of severe infections was observed among patients with persistently higher levels of uric acid but among those with persistently lower levels of albumin. The observed associations were largely consistent across breast and reproductive system cancers, digestive system cancers, and hematological malignancies, supporting the potential utility of these biomarkers as pan-cancer predictors of severe infections after diagnosis.

Our finding on the risk of severe infections align with previous research, which also reported that the risk of sepsis peaked shortly after cancer diagnosis and decreased over time.(6) The observed decreased levels of TC and LDL and increased levels of glucose before cancer diagnosis are also consistent with findings from existing studies.(7, 8) Similarly, increased CRP and IgG levels have also been reported in previous studies.(13) The present study extends therefore the existing knowledgebase by characterizing trajectories of 18 metabolic and inflammatory biomarkers over a time period of 10 years before cancer diagnosis as well as by assessing the associations of such trajectories with risk of severe infections post-cancer diagnosis.

Inflammation is a recognized hallmark of cancer development and progression.(13) Elevated circulating levels of inflammatory biomarkers, such as CRP, have been associated with an increased risk of cancer development in various populations, including the AMORIS cohort.(14-17) Moreover, higher pre-diagnostic levels of haptoglobin and CRP have been linked to an increased risk of premature death among patients with breast cancer.(18) The present study extends this evidence by demonstrating that increasing levels of CRP and IgG during the five years before cancer diagnosis are associated with a heightened risk of severe infections following diagnosis.

Elevated systemic inflammation is also associated with reduced albumin levels,(19) a marker of both inflammatory burden and nutritional status. Consistent with previous findings showing that lower albumin levels are associated with infectious complications during or after cancer treatment,(20-22) our results indicate that persistently low albumin levels during the 10 years preceding cancer diagnosis are associated with an increased risk of severe infections. In addition, elevated uric acid levels have previously been linked to infection severity,(23, 24) a relationship that is extended in the present study by showing that persistently elevated uric acid levels in the decade before cancer diagnosis are associated with a higher risk of severe infections. Taken together, these findings underscore the central role of chronic inflammation and immune dysregulation not only in cancer development but also in patient prognosis and adverse health outcomes, including severe infections, following cancer diagnosis.(25)

Lipids play a crucial role in modulating infection risk in both cancer patients and the general population. Our study showed that persistently high triglyceride levels or low HDL levels were associated with an increased risk of severe infections, consistent with findings in non-cancer populations.(26-28) Cholesterol, particularly LDL and HDL, binds and neutralizes lipopolysaccharides (LPS), protecting against LPS-induced toxicity.(28, 29) Furthermore, lipids facilitate viral replication by regulating membrane trafficking, providing structural components for cellular membranes, and supplying energy for viral replication.(30-32) Dyslipidemia has also been linked to an increased risk of COVID-19 infection among patients with breast cancer.(33)

Hyperglycemia prior to cancer diagnosis may contribute to infection risk by impairing immune function, such as neutrophil activity, and disrupting gastrointestinal and urinary systems.(34) Cancer patients with diabetes are at higher risk of infections, particularly within 90 days after initiating chemotherapy.(35, 36) Moreover, diabetes and hyperglycemia have been associated with increased mortality among cancer patients.(37-39) Prior research has, however, reported that glucose levels within the range of 8.0∼11.5 mmol/L were linked to the lowest sepsis risk among individuals with type 2 diabetes,(40) suggesting that a less stringent glucose control strategy may be appropriate for that population.(41) Interestingly, we found that the risk of severe infection remained largely similar between cancer patients with persistently high (8.0∼11.5 mmol/L) and persistently low (5 mmol/L) glucose levels. However, the observed null association should be interpreted with caution and warrants further confirmation, as only a small number of patients exhibited persistently high glucose levels before cancer diagnosis.

The strengths of our study include the use of large-scale cohort data with repeated biomarker measurements and high-quality follow-up data from nationwide Swedish health registers. Importantly, information on severe infections was collected independently of biomarker measurements. Repeated measurements provide a more comprehensive assessment of a biomarker over time compared to a single snap-shot measurement. Specifically, our results demonstrated that the trajectories of CRP and IgG intersected over the 10-year period and were associated with an increased risk of severe infections after cancer diagnosis. These findings suggest that relying on a single measurement of these biomarkers prior to cancer diagnosis may result in substantial misclassification of patients at elevated risk for severe infections. Furthermore, all blood samples in the AMORIS cohort were analyzed in the same clinical laboratory following well documented and consistently applied standard procedures, ensuring comparability of biomarker measurements between individuals and across different time points. Nevertheless, some limitations should be acknowledged. First, requiring at least three blood samplings before cancer diagnosis may have introduced selection bias. However, no substantial differences were observed in the characteristics of the included and excluded cancer patients. Second, due to limited statistical power, we were unable to analyze the associations between biomarker trajectories and severe infections for all cancer types, although subgroup analyses were performed for the three most common cancer types categorized by organ system. Third, despite adjusting for multiple covariates, residual confounding from unknown or unmeasured confounders such as genetic factors, smoking, body mass index, or alcohol consumption remains possible. Fourth, due to the lack of information, we were unable to investigate whether the observed associations differed by cancer stages according to TNM classification system (data available in the Swedish Cancer Register since 2004) or by cancer treatments. Finally, cancer patients, who died of infections and didn’t have any records of medical visits for severe infections, may be misclassified. Because the Swedish Cause of Death Register generally defines the underlying cause as the disease or injury that initiates the chain of morbid events leading directly to death. However, the misclassification is non-differential in relation to biomarker trajectories and would be expected to attenuate the observed associations.

## Conclusion

We identified distinct trajectories of metabolic and inflammatory biomarkers during the ten years preceding cancer diagnosis. Specific trajectories, such as persistently high levels of triglycerides, haptoglobin, uric acid, persistently low albumin levels, or increasing levels of CRP and IgG, were associated with an elevated risk of severe infections after cancer diagnosis. These findings suggest that longitudinal surveillance of metabolic and inflammatory blood biomarkers before cancer diagnosis could help guide infection prevention and management strategies in cancer patients.

## Data Availability

The data used in this study were obtained from the Swedish National Board of Health and Welfare and Statistics Sweden, which are available for research purposes by researchers who fulfill specific requirements.

## Acknowledgements

The study was supported by the ìShizu Matsumuraîs Donation (grant no. FS-2023:0003) and the Swedish Cancer Society (grant no. 232741Pj). This study was also partially supported by the Initial Founding for High Level Talented Scholars in Nanfang Hospital, Southern Medical University (No. 2023G001) and the Outstanding Youths Development Scheme of Nanfang Hospital, Southern Medical University (Grant No. 2023J009) to QL. DW was supported by the International Postdoc Grant from the Swedish Research Council (grant no. 2023-00206).

## Author contributions

DW, QWL, QW, DZ, and FF designed the study. MF, NH, and FF approved and secured access to the dataset used in this study. DW performed data management and data analysis. QXL, DZ, and DW drafted the manuscript. DZ, QW, QXL, QWL, and DW contributed to interpretation of the results, and all authors reviewed and revised the manuscript. All authors approved the final version for submission.

