## Supplementary materials for "Trajectories of metabolic and inflammatory biomarkers ten years before cancer diagnosis and the risk of subsequent severe infections: a Swedish population-based cohort study"

Table S1. The International Classification of Diseases (ICD) codes used to identify the diagnoses considered in the present study

Table S2. Comparison of characteristics between cancer patients diagnosed during 1985-2005 with three or more blood tests before cancer diagnosis (included in the present study) and those with fewer than three blood tests

Table S3. Parameters of biomarker trajectories from latent class growth modelling

Table S4. Hazard ratios and 95% confidence intervals for the associations of biomarker trajectories with severe infections

Table S5. Hazard ratios and 95% confidence intervals for the associations of biomarker trajectories with severe infections after additionally adjusting for comorbidities

Table S6. Hazard ratios and 95% confidence intervals for the associations between biomarker trajectories and hospitalization for infectious diseases defined by both primary and secondary diagnoses from the Patient Register

Table S7. Hazard ratios and 95% confidence intervals for the associations between biomarker trajectories and hospitalization for infectious diseases stratified by sex

Table S8. Hazard ratios and 95% confidence intervals for the associations between biomarker trajectories and hospitalization for infectious diseases stratified by age at cancer diagnosis

Table S9. Hazard ratios and 95% confidence intervals for the associations between biomarker trajectories and hospitalization for infectious diseases stratified by a history of severe infections during the 10 years before cancer diagnosis

Table S10. Hazard ratios and 95% confidence intervals for the associations between biomarker trajectories and hospitalization for infectious diseases among patients with breast and reproductive system cancers, digestive system cancers, and hematological malignancies, respectively

Figure S1. Crude hazard ratios and 95% confidence intervals of hospitalization for infectious diseases in relation to biomarker trajectories

Figure S2. Hazards of sepsis diagnosis after a cancer diagnosis in relation to biomarker trajectories

**Table S1. The International Classification of Diseases (ICD) codes used to identify the diagnoses considered in the present study**

| **Diagnoses** | **ICD-7** | **ICD-8** | **ICD-9** | **ICD-10** |
| --- | --- | --- | --- | --- |
| Cancer | 140-207 | - | - | - |
| Infection | - | **Bacterial infection：**000.01-005.99, 008.00-008.30, 010.99-018.98, 020.00-039.98, 073.99, 076.99, 079.30, 080.99-083.99, 088.99-104.98, 320.00-320.80, 322.00-322.03, 361.00-361.09, 362.02, 366.00, 369.00, 380.00-380.01, 382.00-383.99, 390.97-392.99, 421.00, 461.00-461.09, 462.02, 463.01, 481,99-482.98, 501.99, 508.00-508.02, 510.01-510.09, 511.10, 513.99, 522.50, 527.30, 528.00, 528.30, 540.03, 562.00-562.19, 566.00-566.01, 567.00-567.02, 569.00, 577.01, 590.00-590.99, 595.00-595.02, 597.00, 599.02, 611.00, 611.01, 612.01-614.99, 616.00-616.03, 620.00-620.99, 622.00-622.19, 629.40, 630.00-630.09, 635.00-636.09, 645.90-645.91, 670.00-670.09, 678.02, 680.00-682.99, 684.00-684.09, 710.00-710.09, 720.00-720.29, 732.99, 761,00, 763.00, 998.50, 999.30  **Virus infection:**  008.80-008.98, 040.00-043.99, 045.00-065.99, 067.00-072.09, 074.00-075.09, 078.00-079.20, 079.40-079.99, 099.92, 460.99, 464.01-480,99, 508.03, 761.20, 761.30  **Other infection:**  006.00-007.99, 009.00-009.98, 084.00-087.99, 099.96-099.99, 110.00-130.10, 130.99-131.99, 136.09, 320.88-320.99, 360.00, 380.02-381.99, 384.00-384.08, 420.00-420.09, 421.98, 422.97-422.99, 462.01, 462.09, 463.09, 466.99, 483.99-486.09, 503.00-503.09, 540.00-540.02, 540.04-540.99, 572,99, 686.00-686.98, 761,40, 763.10, 763.98, 778,60 | **Bacterial infection：**001-005X, 008A-F, 010-041X, 073, 076, 078D, J, 790H, 080-083X, 087-099D, 100-104, 245A, 254B, 320-X, 324-X, 360A, 373B, 375D, 376A, 382A-E, 383A-X, 390-392X, 421A, 461-X, 475, 481-482X, 510-X, 511B, 513-B, 522E, H, 526E, 527D, 528A, D, 540B, 562-B, 566, 567-C, 569F, 575A, 590-X, 597A, 595-D, X, 597W, 599A, 611A, 614-F, W-X, 615A, X, 616-X, 634A, 635A, 636A, 637A, 638A, 639A, 646F, G, 647A, B, D, 658E, 659D, 670, 675-B, W-X, 681-686X, 711A, E, 728A, 729E, 730-D, X, 771D, 996G, 998F, 999D  **Virus infection:**  008H-M, 045-066, 070-072X, 074-075, 077-078H, 078W-079X, 279K, 321B-H, 323A, 323C-D, 460, 464-465X, 480-X, 487-W, 647F, G, 711F, 771A, B, 790W  **Other infection:**  006-007X, 008W, 009-D, 084-086X, 099E-X, 110-136X, 321A, 321W, 370E-F, X, 372A-D, 380B, C, 381A, 382X, 420- 422X, 462-463, 466-B, 473-X, 483, 485-486, 490, 491B, 540A, X, 572A, 647C, E, W, X, 680A, 711G-X, 727A, 770A, 771C, E-W | **Bacterial infection：**  A00-05.9, A15-17.9, A20-28.9, A30 -58, A65 -79.9, B95-96.8, E06.0, E32.1, G00-00.9, G01, G04.2, G05.0, G06-06.2*, G07, H00.0, H01.0, H04.3, H05.0, H44.0, H60.0-60.1, H66.0-66.4, H70.0-70.9, I00-02.9, J01-01.9*, J02.0, J03.0, J13-15.9, J16.0, J20.0-20.2, J34.0, J36*, J39.0-39.1, J85.1-85.3, J86-86.9*, K04.6-04.7, K05.2, K11.3, K12.2, K14.0, K35.1, K57-57.9, K61-61.4, K63.0, K65.0*, K81.0, K85, L00 -08.9, M00-00.9, M46.3*, M60.0*, M86-86.9*, N10-12*, N13.6*, N15.1, N15.9, N30-30.3*, N30.8-30.9*, N34-34.1*, N39.0*, , N61, N70-76.8*, N98.0, O07.0, O07.5, O08.0, O23-23.9, O41.1, O75.3, O85-86.8*, O91-91.1, O98.0-98.2, P23.1-23.6, P36, P37.0, T80.2, T81.4, T82.6-82.7, T83.5-83.6, T84.5-84.7, T85.7, T88.0, Z22.0-22.3  **Virus infection:**  A08-08.4, A60-60.9, A63.0, A80-89, A90-99, B00-06.0, B06.8-09, B15-19.9, B20-24, B25-34, B97-97.8, G02.0, G05.1, J00, J04-06.9*, J10-11.8, J12-12.9, J20.3-20.7, J21.0, O35.3, O98.4-98.5, P23.0, P35, Z21, Z22.5-22.6  **Other infection:**  A06-07.9, A08.5, A09, A59-59.9, A63, A63.8-64, B35-49, B50 -89, B99, G02.1-02.8, G04, G04.9, G05.2, H10.0, H10.3-10.9, H16.2-16.3, H16.9, H32, H60, H60.3, H65.0-65.1, H66.9, I30.0-30.9, I33.0-33.9, I40.0, J02*, J02.8-02.9*, J03*, J03.8-03.9*, J16, J16.8, J18-18.9, J20, J20.8-21, J21.8-21.9, J22, J32-32.9*, J35.0, J37-37.1*, J40-42, K35, K35.9, K75.0, L30.3, M46.5, M65.1, M71.1, O98.3, O98.6-98.9, P23.8-23.9, P37.1-39.9, Z22.4, Z22.8-22.9 |
| Sepsis | - | - | 036C, 036D, 036E, 036X, 038, 084, 112F, 117D, 286G, 999D | A02.1, A04.0, A04.1, A04.2, A04.3, A39, A40, A41, A42.7, A48, A90-A99, B37.7, B38.7, B39.3, B40.7, B41.7, B42.7, B44.7, B45.7, B46.4, B95-99, D65, T80.2 |
| Psychiatric disorders | - | 290-315 | 290-319 | F00-F99 |
| Diabetes | - | 250 | 250 | E10-E14 |

**Table S2. Comparison of characteristics between cancer patients diagnosed during 1985-2005 with three or more blood tests before cancer diagnosis (included in the present study) and those with fewer than three blood tests**

| **Characteristics** | **Cancer patients diagnosed during 1985-2005 with fewer than three blood tests before cancer diagnosis** | **Cancer patients diagnosed during 1985-2005 with three or more blood tests before cancer diagnosis (included in the present study)** |
| --- | --- | --- |
|  | **(N=62,902)** | **(N=10,837)** |
| Sex: male, N (%) | 33,090 (52.6) | 5476 (50.5) |
| Age at cancer diagnosis, years/mean (SD) | 63.4 (13.8) | 65.9 (11.7) |
| Year of diagnosis, N (%) |  |  |
| 1985-1995 | 23,543 (37.4) | 4185 (38.6) |
| 1996-2005 | 39,359 (62.6) | 6652 (61.4) |
| Country of birth, N (%) |  |  |
| Sweden | 55,246 (87.8) | 9385 (86.6) |
| Outside Sweden | 7656 (12.2) | 1452 (13.4) |
| Education at the recruitment, N (%) |  |  |
| Primary | 15,624 (24.8) | 2523 (23.3) |
| Secondary | 28,464 (45.3) | 5060 (46.7) |
| Tertiary | 13,788 (21.9) | 2456 (22.7) |
| Unknown | 5026 (8.0) | 798 (7.4) |
| Employment status at the recruitment, N (%) |  |  |
| Non-gainfully employed or unemployed | 17,231 (27.4) | 2913 (26.9) |
| Gainfully employed | 44,803 (71.2) | 7867 (72.6) |
| Unknown | 868 (1.4) | 57 (0.5) |
| Income at the recruitment, N (%) |  |  |
| Below 1st tertile | 20,597 (32.7) | 3768 (34.8) |
| 1st-2nd tertile | 20,259 (32.2) | 3196 (29.5) |
| Above 2nd tertile | 21,320 (33.9) | 3849 (35.5) |
| Unknown | 726 (1.2) | 24 (0.2) |
| Severe infections during the 10 years before cancer diagnosis: yes, N (%)* | 7394 (11.8) | 1346 (12.4) |
| Psychiatric disorders before cancer diagnosis: yes, N (%) | 5105 (8.1) | 907 (8.4) |
| Diabetes before cancer diagnosis: yes, N (%) | 3128 (5.0) | 724 (6.7) |

SD=standard deviation.

* Severe infections included hospitalization for infectious diseases and any diagnosis of sepsis at a hospital visit.

**Table S3. Parameters of biomarker trajectories from latent class growth modelling**

| **Biomarkers** | | **C** | **Fixed effect** | | | **Random intercept** | | | **Random intercept and slope** | | |
| --- | --- | --- | --- | --- | --- | --- | --- | --- | --- | --- | --- |
|  |  |  | **Logit** | **BIC** | **Class membership (%)** | **Logit** | **BIC** | **Class membership (%)** | **Logit** | **BIC** | **Class membership (%)** |
| Glucose | L^a^ | 2 | 9145.56 | -18237.14 | 91.98/8.02 | **13487.03** | **-26911.08** | **93.18/6.82** | 11713.05 | -23345.13 | 57/43 |
|  |  | 3 | 11715.03 | -23349.09 | 84.47/11.01/4.52 | 14082.05 | -28074.14 | 2.86/92.29/4.85 | 13643.33 | -27178.7 | 93.15/0/6.85 |
|  |  | 4 | 12983.14 | -25858.31 | 5.3/15.93/76.07/2.7 | 14082.05 | -28047.15 | 0/92.16/4.94/2.9 | 13643.33 | -27151.71 | 0/92.66/0/7.34 |
|  |  | 5 | 13455.61 | -26776.26 | 5.13/2.98/70.84/19.72/1.32 | 14082.05 | -28020.16 | 3.01/0/0/91.41/5.58 | 13643.33 | -27151.71 | 0/92.66/0/7.34 |
|  | Q^b^ | 2 | 9146.66 | -18221.35 | 91.96/8.04 | 13488.23 | -26895.5 | 93.15/6.85 | 13655.11 | -27211.25 | 6.6/93.4 |
|  |  | 3 | 11721.25 | -23334.53 | 10.77/84.77/4.46 | 14237 | -28357.04 | 3.12/91.62/5.26 | 13655.11 | -27175.26 | 6.72/93.28/0 |
|  |  | 4 | 12986.49 | -25829.04 | 75.99/5.27/16.03/2.7 | 14536.16 | -28919.37 | 91.99/2.7/2.96/2.35 | 13655.11 | -27139.28 | 7.09/92.91/0/0 |
|  |  | 5 | 13470.22 | -26760.49 | 5.14/2.91/19.78/70.76/1.41 | 14746.5 | -29304.06 | 2.45/4.51/90.57/0.94/1.54 | 13655.11 | -27103.29 | 7.28/92.72/0/0/0 |
| Fructosamine | L | 2 | 23352.45 | -46652.4 | 91.13/8.87 | **27215.95** | **-54370.65** | **94.99/5.01** | 26694.62 | -53310.48 | 54.45/45.55 |
|  |  | 3 | 25318.66 | -50558.58 | 53.58/40.84/5.58 | 27496.49 | -54905.48 | 3.39/94.6/2.01 | 27359.82 | -54614.64 | 0/95.18/4.82 |
|  |  | 4 | 26278.81 | -52452.63 | 55.63/34.39/7.19/2.79 | 27558.02 | -55002.29 | 3.25/1.51/94.41/0.84 | 27359.82 | -54588.39 | 94.58/0/0/5.42 |
|  |  | 5 | 26690.51 | -53249.78 | 13.74/57.23/5.04/22.03/1.97 | 27640.5 | -55140.99 | 3.03/0.44/94.25/1.52/0.76 | * | * | * |
|  | Q | 2 | 23355.41 | -46640.81 | 91.2/8.8 | 27219.48 | -54360.21 | 95.01/4.99 | 27381.73 | -54667.21 | 4.03/95.97 |
|  |  | 3 | 25319.79 | -50534.59 | 53.63/40.76/5.61 | 27541.5 | -54969.25 | 3.28/94.72/2 | 27649.27 | -55167.29 | 3.33/95.13/1.54 |
|  |  | 4 | 26286.22 | -52432.45 | 34.61/7.12/55.53/2.74 | 27640.17 | -55131.59 | 3.04/94.33/0.95/1.68 | 27768.2 | -55370.15 | 3/0.7/94.93/1.38 |
|  |  | 5 | 26698.8 | -53222.59 | 12.79/23.65/56.5/4.96/2.11 | 27824.91 | -55466.07 | 1.17/94.1/0.1/2.92/1.71 | 27894.47 | -55587.7 | 1.87/0.14/94.55/2.31/1.13 |
| TC | L | 2 | 18155.41 | -36256.35 | 41.5/58.5 | 26198.67 | -52333.79 | 4.13/95.87 | 26160.37 | -52239.02 | 48.24/51.76 |
|  |  | 3 | 21924.78 | -43767.85 | 17.93/48.82/33.25 | 26238.42 | -52386.05 | 2.09/94.6/3.32 | 26252.71 | -52396.47 | 83.34/0.39/16.27 |
|  |  | 4 | 23747.22 | -47385.49 | 9.08/42.04/32.16/16.73 | 26327.88 | -52537.72 | 95.59/3.47/0.6/0.34 | 26326.95 | -52517.72 | 1.7/2.15/96.15/0 |
|  |  | 5 | 24728.59 | -49321 | 3.27/17.98/33.84/33.23/11.67 | 26368.41 | -52591.56 | 0.58/3.5/1.87/93.72/0.33 | * | * | * |
|  | Q | 2 | **18168.22** | **-36263.8** | **41.55/58.45** | 26225.83 | -52369.96 | 4.34/95.66 | 26350.2 | -52600.53 | 1.68/98.32 |
|  |  | 3 | 21941.86 | -43774.78 | 17.93/48.85/33.21 | 26393.63 | -52669.23 | 3.75/95.1/1.15 | 26486.2 | -52836.2 | 1.63/97.4/0.97 |
|  |  | 4 | 23770.64 | -47396 | 9.09/32.28/42.1/16.53 | 26463.71 | -52773.07 | 1.53/95.02/3.1/0.35 | 26539.78 | -52907.06 | 1.61/96.32/0.79/1.29 |
|  |  | 5 | 24747.73 | -49313.88 | 3.18/17.5/33.61/33.82/11.89 | 26540.24 | -52889.81 | 0.32/3.39/1.4/94.65/0.24 | 26558.96 | -52909.11 | 1.65/86.66/10.57/0.78/0.34 |
| Triglycerides | L | 2 | -27188.69 | 54431.82 | 60.94/39.06 | -21198.79 | 42461.09 | 4.01/95.99 | -21178.98 | 42439.62 | 51.23/48.77 |
|  |  | 3 | -23829.1 | 47739.87 | 37.73/46.46/15.81 | -21098.39 | 42287.52 | 4.68/92.59/2.73 | -21098.41 | 42305.7 | 14.13/85.87/0 |
|  |  | 4 | -22462.16 | 45033.2 | 23.32/28.38/40.27/8.04 | -21062.29 | 42242.53 | 3.51/7.09/87.45/1.96 | -21098.41 | 42332.92 | 23.66/0/76.34/0 |
|  |  | 5 | -21870.07 | 43876.24 | 16.23/33.97/31.06/15.05/3.68 | -21011.17 | 42167.52 | 0.13/85.5/4.48/1.88/8.01 | * | * | * |
|  | Q | 2 | -27187.71 | 54448 | 60.88/39.12 | -21193.26 | 42468.19 | 4.57/95.43 | **-21081.08** | **42261.97** | **13.75/86.25** |
|  |  | 3 | -23827.6 | 47764.08 | 37.66/46.5/15.83 | -21088.33 | 42294.62 | 6.43/90.35/3.22 | -21020.15 | 42176.4 | 1.05/13.87/85.07 |
|  |  | 4 | -22454.22 | 45053.61 | 28.28/40.3/23.4/8.01 | -21040.76 | 42235.77 | 4.09/8.32/85.21/2.37 | -20997.82 | 42168.03 | 0.91/10.59/87.02/1.48 |
|  |  | 5 | -21845.4 | 43872.27 | 16.56/33.91/31.05/14.84/3.65 | -20979.1 | 42148.75 | 5.33/0.09/85.15/2.18/7.25 | -20959.86 | 42128.41 | 0.13/1.96/0.24/84.87/12.81 |
| LDL | L | 2 | 325.58 | -605.65 | 30.07/69.93 | 1637.76 | -3222.43 | 4.06/95.94 | 1605.76 | -3143.25 | 47.94/52.06 |
|  |  | 3 | 954.46 | -1840.65 | 12.14/43.78/44.08 | 1682.48 | -3289.11 | 7.57/88.52/3.91 | 1657.61 | -3224.19 | 87.76/12.24/0 |
|  |  | 4 | **1301.58** | **-2512.14** | **6.75/22.7/46.93/23.62** | 1682.48 | -3266.35 | 4.72/8.48/86.8/0 | 1657.61 | -3201.43 | 85.27/14.73/0/0 |
|  |  | 5 | 1417.11 | -2720.43 | 5.64/17.32/29.2/34.69/13.15 | 1682.48 | -3243.59 | 7.31/0/0/81.51/11.17 | * | * | * |
|  | Q | 2 | 327.67 | -594.66 | 30.12/69.88 | 1639.26 | -3210.26 | 96.29/3.71 | 1658.15 | -3232.86 | 90.45/9.55 |
|  |  | 3 | 953.08 | -1815.14 | 15.39/37.43/47.18 | 1685.99 | -3273.38 | 7.57/89.18/3.25 | 1697.96 | -3282.15 | 1.12/90.35/8.53 |
|  |  | 4 | 1302.52 | -2483.68 | 6.81/22.6/47.03/23.57 | 1721.35 | -3313.75 | 4.42/88.17/4.82/2.59 | 1724.28 | -3304.43 | 1.52/89.03/3.05/6.4 |
|  |  | 5 | 1417.96 | -2684.22 | 5.69/17.37/34.64/29.15/13.15 | 1734.89 | -3310.5 | 0.46/5.08/3/4.16/87.3 | 1724.28 | -3274.09 | 1.88/7.47/0/87.46/3.2 |
| HDL | L | 2 | 1219.77 | -2394.46 | 46.99/53.01 | 2408.18 | -4763.78 | 87.81/12.19 | 2440.66 | -4813.7 | 44.32/55.68 |
|  |  | 3 | 1995.93 | -3924.25 | 27.16/42.9/29.95 | 2432.99 | -4790.86 | 26.12/57.1/16.78 | 2445.66 | -4801.17 | 47.27/30.27/22.46 |
|  |  | 4 | 2202 | -4313.86 | 18.09/29.34/34.15/18.42 | 2445.55 | -4793.45 | 0.87/34.04/57.21/7.87 | 2448.59 | -4784.5 | 24.92/3.5/48.31/23.28 |
|  |  | 5 | 2355.11 | -4597.55 | 13.11/27.21/31.09/21.91/6.67 | 2450.13 | -4780.07 | 0.93/21.31/45.36/22.68/9.73 | * | * | * |
|  | Q | 2 | 1224.02 | -2387.94 | 47.1/52.9 | 2411.87 | -4756.14 | 88.47/11.53 | **2451.88** | **-4821.12** | **57.32/42.68** |
|  |  | 3 | 1999.81 | -3909.47 | 27.05/42.95/30 | 2436.18 | -4774.7 | 1.2/94.81/3.99 | 2460.44 | -4808.19 | 24.04/46.01/29.95 |
|  |  | 4 | 2208.45 | -4296.71 | 18.63/33.55/30.16/17.65 | 2462.76 | -4797.82 | 1.53/56.5/9.4/32.57 | 2466.71 | -4790.69 | 1.64/57.27/5.85/35.25 |
|  |  | 5 | 2359.59 | -4568.93 | 12.95/21.69/27.1/31.48/6.78 | 2471.93 | -4786.12 | 45.25/1.26/22.68/20.6/10.22 | 2474.85 | -4776.93 | 1.31/23.88/45.68/7.21/21.91 |
| LDL/HDL ratio | L | 2 | -2645.02 | 5335.11 | 38.49/61.51 | **-1448.23** | **2949.03** | **5.25/94.75** | -1455.09 | 2977.78 | 48.33/51.67 |
|  |  | 3 | -2021.79 | 4111.18 | 20.56/49.75/29.69 | -1432.26 | 2939.64 | 4.65/81.36/14 | -1426.24 | 2942.61 | 5.3/16.13/78.57 |
|  |  | 4 | -1711.79 | 3513.73 | 7.76/29.69/45.71/16.84 | -1426.23 | 2950.12 | 2.24/79.5/2.52/15.75 | -1417.93 | 2948.54 | 5.41/0.82/75.94/17.82 |
|  |  | 5 | -1590 | 3292.68 | 4.92/16.18/28.59/37.51/12.79 | -1415.36 | 2950.9 | 16.13/2.13/0.6/79/2.13 | * | * | * |
|  | Q | 2 | -2640.75 | 5341.6 | 38.38/61.62 | -1444.78 | 2957.16 | 95.02/4.98 | -1429.96 | 2942.54 | 1.97/98.03 |
|  |  | 3 | -2014.13 | 4118.41 | 20.61/49.92/29.47 | -1428.17 | 2953.99 | 4.16/83.93/11.92 | -1408.38 | 2929.43 | 2.35/94.53/3.12 |
|  |  | 4 | -1707.93 | 3536.05 | 7.71/29.52/45.76/17 | -1404.15 | 2936 | 3.01/85.07/10.22/1.69 | -1408.38 | 2959.47 | 2.73/92.51/0/4.76 |
|  |  | 5 | -1585.66 | 3321.55 | 4.92/28.54/16.35/37.45/12.74 | -1385.1 | 2927.95 | 3.17/79.44/15.97/0.22/1.2 | -1379.31 | 2931.39 | 5.47/74.03/19.03/0.22/1.26 |
| ApoA1 | L | 2 | 5529.91 | -11014.26 | 52.82/47.18 | 6672.01 | -13290.86 | 2.72/97.28 | 6674.26 | -13280.18 | 50.5/49.5 |
|  |  | 3 | 6179.02 | -12289.69 | 51.01/22.91/26.08 | 6699.01 | -13322.08 | 2.67/97.08/0.25 | 6674.27 | -13257.41 | 28.45/42.6/28.95 |
|  |  | 4 | 6421.23 | -12751.33 | 8.06/39.38/35.2/17.37 | 6703.18 | -13307.65 | 1.81/0.5/97.43/0.25 | 6704.79 | -13295.68 | 2.11/96.78/0.25/0.86 |
|  |  | 5 | 6549.25 | -12984.59 | 6.34/36/28.65/24.12/4.88 | 6711.21 | -13300.91 | 0.2/0.45/1.66/97.43/0.25 | * | * | * |
|  | Q | 2 | 5533.51 | -11006.27 | 52.72/47.28 | **6680** | **-13291.66** | **49.9/50.1** | 6710.09 | -13336.64 | 99.04/0.96 |
|  |  | 3 | 6181.62 | -12272.11 | 50.65/22.96/26.38 | 6720.76 | -13342.8 | 1.91/97.38/0.7 | 6730.99 | -13348.08 | 0.76/98.49/0.76 |
|  |  | 4 | 6424.13 | -12726.75 | 8.06/35.15/39.53/17.27 | 6726.34 | -13323.58 | 1.71/1.36/96.12/0.81 | 6759.86 | -13375.44 | 0.76/98.14/0.65/0.45 |
|  |  | 5 | 6551.97 | -12952.06 | 6.19/24.02/28.45/36.4/4.93 | 6761.75 | -13364.02 | 0.65/95.97/1.01/1.91/0.45 | 6765.09 | -13355.51 | 0.6/0.81/1.16/96.98/0.45 |
| ApoB | L | 2 | 379.46 | -713.6 | 40.69/59.31 | **1728.13** | **-3403.39** | **5.72/94.28** | 1745.73 | -3423.47 | 51.02/48.98 |
|  |  | 3 | 1080.19 | -2092.39 | 15.36/47.98/36.65 | 1754.82 | -3434.1 | 2.25/93.76/3.99 | 1784.85 | -3479.07 | 93.5/6.29/0.21 |
|  |  | 4 | 1368.83 | -2647.03 | 10.17/33.56/41.11/15.15 | 1779.75 | -3461.32 | 1.89/7.18/84.22/6.71 | 1792.96 | -3472.62 | 0.21/9.7/89.88/0.21 |
|  |  | 5 | 1513.7 | -2914.1 | 4.82/18.46/35.08/33.14/8.5 | 1781.88 | -3442.91 | 1.05/0.79/85.32/7.08/5.77 | * | * | * |
|  | Q | 2 | 384.92 | -709.41 | 40.69/59.31 | 1729.88 | -3391.79 | 4.82/95.18 | 1763.72 | -3444.36 | 1.05/98.95 |
|  |  | 3 | 1081.66 | -2072.68 | 15.26/47.93/36.81 | 1758.15 | -3418.1 | 1.89/94.13/3.99 | 1793.24 | -3473.17 | 1.26/6.14/92.61 |
|  |  | 4 | 1371.11 | -2621.37 | 10.07/33.3/41.37/15.26 | 1771.17 | -3413.93 | 1.68/1.57/93.55/3.2 | 1800.41 | -3457.3 | 1.26/89.41/2.05/7.29 |
|  |  | 5 | 1522.83 | -2894.59 | 1.21/12.85/37.28/35.34/13.32 | 1800.35 | -3442.08 | 1.36/77.92/8.34/5.61/6.76 | 1801.76 | -3429.8 | 1.31/10.44/80.55/6.82/0.89 |
| ApoB/ApoA1 ratio | L | 2 | -964.18 | 1973.31 | 36.01/63.99 | **620.75** | **-1189.06** | **10.14/89.86** | 630.38 | -1193.34 | 49.61/50.39 |
|  |  | 3 | -226.69 | 520.8 | 22.3/50.5/27.2 | 635.45 | -1195.97 | 3.9/94.7/1.39 | 658.47 | -1227.03 | 1.73/7.97/90.3 |
|  |  | 4 | 208.5 | -327.09 | 7.41/46.21/27.2/19.18 | 643.6 | -1189.79 | 10.14/2.4/86.29/1.17 | 661.1 | -1209.82 | 2.84/8.08/88.74/0.33 |
|  |  | 5 | 369.97 | -627.56 | 6.24/21.13/28.54/33.78/10.31 | 662.55 | -1205.23 | 1.95/80.04/0.56/8.58/8.86 | * | * | * |
|  | Q | 2 | -960.51 | 1980.96 | 36.23/63.77 | 626.6 | -1185.77 | 6.02/93.98 | 650.09 | -1217.76 | 1.45/98.55 |
|  |  | 3 | -220.88 | 531.67 | 22.3/50.72/26.98 | 642.86 | -1188.32 | 3.68/88.35/7.97 | 675.15 | -1237.91 | 1.78/89.97/8.25 |
|  |  | 4 | 211.78 | -303.69 | 7.47/27.03/19.34/46.15 | 658.54 | -1189.72 | 4.24/10.87/82.27/2.62 | 685.06 | -1227.77 | 1.95/0.22/89.58/8.25 |
|  |  | 5 | 372.22 | -594.59 | 6.24/20.9/33.84/28.71/10.31 | 679.96 | -1202.58 | 3.9/26.42/3.12/56.47/10.09 | 686.48 | -1200.65 | 1.56/87.96/0.84/0.45/9.2 |
| Hemoglobin | L | 2 | 19063.53 | -38077.9 | 37.96/62.04 | 23484.78 | -46912.21 | 3.45/96.55 | 23312.21 | -46550.68 | 44.64/55.36 |
|  |  | 3 | 20996.83 | -41919.91 | 51.44/37.93/10.64 | 23575.05 | -47068.15 | 3.48/95.55/0.97 | 23637.58 | -47176.82 | 2.15/97.85/0 |
|  |  | 4 | 21858.19 | -43618.05 | 3.78/48.37/25.55/22.29 | 23781.72 | -47456.92 | 0.94/3.37/94.67/1.02 | 23637.58 | -47152.24 | 2.35/97.65/0/0 |
|  |  | 5 | 22407.28 | -44691.65 | 1.57/39.17/31.77/18.09/9.39 | 23814.02 | -47496.94 | 0.91/0.88/94.34/0.5/3.37 | * | * | * |
|  | Q | 2 | 19201.39 | -38337.24 | 35.44/64.56 | 23768.45 | -47463.15 | 2.96/97.04 | 23946.13 | -47802.13 | 2.57/97.43 |
|  |  | 3 | 21269.23 | -42440.14 | 11.93/51.13/36.93 | 23900.13 | -47693.73 | 2.98/95.58/1.44 | 24061.21 | -47999.51 | 2.6/96.24/1.16 |
|  |  | 4 | **22090.41** | **-44049.71** | **6.74/44.72/26.71/21.82** | 24111.98 | -48084.65 | 1.57/3.18/93.78/1.46 | 24160.15 | -48164.61 | 1.6/94.7/2.68/1.02 |
|  |  | 5 | 22635.28 | -45106.68 | 1.91/14.45/11.05/36.38/36.22 | 24145.78 | -48119.47 | 1.57/92.76/2.79/1.44/1.44 | 24235.68 | -48282.9 | 0.61/94.17/2.21/1.77/1.24 |
| Globulin | L | 2 | **5236.87** | **-10428.79** | **67.13/32.87** | 6139.21 | -12225.97 | 98.21/1.79 | 6141.32 | -12215.22 | 51.34/48.66 |
|  |  | 3 | 5733.65 | -11399.88 | 43.64/52.06/4.3 | 6168.65 | -12262.39 | 38.39/60.44/1.17 | 6174.03 | -12258.18 | 99.11/0/0.89 |
|  |  | 4 | 5965.64 | -11841.39 | 21.37/51.34/24.22/3.07 | 6174.83 | -12252.27 | 1/80.64/17.19/1.17 | 6174.03 | -12235.7 | 0/0/98.72/1.28 |
|  |  | 5 | 6057.66 | -12002.95 | 13.5/39.9/34.93/10.04/1.62 | 6194.25 | -12268.65 | 1/0.17/20.7/77.06/1.06 | * | * | * |
|  | Q | 2 | 5243.41 | -10426.89 | 66.57/33.43 | 6145.42 | -12223.42 | 98.38/1.62 | 6171.52 | -12260.65 | 99.27/0.73 |
|  |  | 3 | 5740.37 | -11390.86 | 43.81/51.9/4.3 | 6169.11 | -12240.84 | 96.93/1.45/1.62 | 6205.23 | -12298.09 | 1.28/97.27/1.45 |
|  |  | 4 | 5975.95 | -11832.04 | 20.87/51.34/24.78/3.01 | 6200.02 | -12272.69 | 29.35/1.62/67.91/1.12 | 6219.72 | -12297.11 | 1.06/28.01/69.81/1.12 |
|  |  | 5 | 6062.94 | -11976.05 | 13.45/10.44/39.62/34.6/1.9 | 6209.57 | -12261.84 | 21.15/0.39/2.68/73.05/2.73 | 6227.65 | -12283.01 | 0.33/45.7/50.33/2.46/1.17 |
| Haptoglobin | L | 2 | -2359.56 | 4770.48 | 40.52/59.48 | 926.02 | -1792.12 | 97.43/2.57 | 845.25 | -1613.48 | 52.28/47.72 |
|  |  | 3 | **-803.81** | **1684.65** | **15.34/58.02/26.64** | 1006.2 | -1926.81 | 0.92/96.51/2.57 | 900.63 | -1698.56 | 0/97.31/2.69 |
|  |  | 4 | -53.8 | 210.31 | 6.24/34.99/47.83/10.94 | 1047.23 | -1983.2 | 1.8/96.3/0.88/1.02 | 908.7 | -1689.03 | 95.91/0.31/0/3.78 |
|  |  | 5 | 356.34 | -584.3 | 3.24/19.17/30.25/43.53/3.8 | 1084.13 | -2031.33 | 95.07/1.32/1.92/0.88/0.81 | * | * | * |
|  | Q | 2 | -2302.18 | 4672.82 | 40.98/59.02 | 1035.8 | -1994.57 | 96.97/3.03 | 1118.53 | -2142.91 | 97.64/2.36 |
|  |  | 3 | -732.72 | 1568.14 | 15.66/26.47/57.87 | 1128.77 | -2146.28 | 3.01/96.31/0.67 | 1166.1 | -2203.82 | 95.6/2.23/2.17 |
|  |  | 4 | 10.44 | 116.05 | 35.57/47.2/7.22/10.02 | 1215.13 | -2284.78 | 1/95.45/0.61/2.94 | 1248.66 | -2334.71 | 96.03/2.53/0.52/0.92 |
|  |  | 5 | 424.45 | -677.73 | 3.15/18.43/31.34/42.73/4.36 | 1257.38 | -2335.03 | 1.09/0.54/2.65/94.51/1.21 | 1274.82 | -2352.8 | 0.54/2.57/0.81/95.01/1.07 |
| CRP | L | 2 | -23345.06 | 46740 | 5.42/94.58 | -23079.27 | 46216.73 | 87.3/12.7 | -23065.54 | 46205.9 | 55.16/44.84 |
|  |  | 3 | -23180.63 | 46436.09 | 1.2/90.71/8.09 | -22913.93 | 45911 | 2.97/84.48/12.55 | -22894.21 | 45888.19 | 95/1.77/3.24 |
|  |  | 4 | -23015.59 | 46130.94 | 3.53/80.07/9.61/6.79 | -22877.7 | 45863.48 | 0.96/84.8/11.18/3.06 | -22863.63 | 45851.97 | 90.88/4.63/3.63/0.86 |
|  |  | 5 | -22905.57 | 45935.85 | 0.47/4.66/79.9/10.69/4.29 | -22845.44 | 45823.9 | 0.51/83.08/11.65/1.86/2.89 | * | * | * |
|  | Q | 2 | -23312.57 | 46691.66 | 94.21/5.79 | **-23049.26** | **46173.34** | **11.99/88.01** | -22876.14 | 45843.72 | 91.64/8.36 |
|  |  | 3 | -23066.08 | 46231.93 | 82.86/13.51/3.63 | -22895.97 | 45900.02 | 15.03/81.49/3.48 | -22671.08 | 45466.86 | 2.97/87.2/9.83 |
|  |  | 4 | -22909.81 | 45952.63 | 10.98/79.19/5.76/4.07 | -22744.36 | 45630.05 | 8.16/85.63/3.48/2.72 | -22616.7 | 45391.35 | 2.99/85.27/9.98/1.77 |
|  |  | 5 | -22764.63 | 45695.52 | 10.35/9.88/4.17/74.41/1.2 | -22651.53 | 45477.65 | 2.77/17.75/73.28/4.49/1.72 | -22584.85 | 45360.91 | 2.13/2.65/9.71/83.67/1.84 |
| IgG | L | 2 | -138.82 | 315.33 | 68.16/31.84 | **575.99** | **-1108.02** | **94.38/5.62** | 581.18 | -1105.84 | 52.81/47.19 |
|  |  | 3 | 148.52 | -240.52 | 53.93/36.33/9.74 | 585.09 | -1107.38 | 0.75/93.82/5.43 | 611.17 | -1146.97 | 97.38/0/2.62 |
|  |  | 4 | 320.6 | -565.83 | 26.03/22.28/47.75/3.93 | 612.61 | -1143.57 | 0.75/6.74/89.7/2.81 | * | * | * |
|  |  | 5 | 428.29 | -762.37 | 9.93/17.6/32.96/35.77/3.75 | 612.61 | -1124.73 | 1.31/0/85.02/10.67/3 | * | * | * |
|  | Q | 2 | -137.08 | 324.4 | 68.16/31.84 | 582.02 | -1107.52 | 94.94/5.06 | 614.7 | -1160.32 | 98.13/1.87 |
|  |  | 3 | 153.24 | -231.12 | 36.7/53.56/9.74 | 612.37 | -1143.09 | 6.74/92.13/1.12 | 631.21 | -1168.22 | 2.62/95.51/1.87 |
|  |  | 4 | 327.94 | -555.39 | 26.22/47.57/22.1/4.12 | 606.7 | -1106.64 | 5.99/89.51/1.87/2.62 | 632.83 | -1146.34 | 3/3.37/91.76/1.87 |
|  |  | 5 | 432.37 | -739.13 | 10.67/32.4/18.16/34.64/4.12 | 625.79 | -1119.69 | 2.06/3.18/89.33/3.56/1.87 | * | * | * |
| Leukocyte | L | 2 | -2150.47 | 4349.73 | 69.58/30.42 | 1952.33 | -3847.74 | Feb-98 | 1849.88 | -3626.57 | 52.31/47.69 |
|  |  | 3 | **-507.54** | **1088.26** | **47.48/45.51/7** | 1985.79 | -3890.28 | 3.32/94.7/1.97 | 2006.87 | -3916.16 | 98.23/0/1.77 |
|  |  | 4 | 547.93 | -998.29 | 28.66/50.28/19.33/1.74 | 2040.52 | -3975.33 | 96.26/2.03/1.09/0.62 | * | * | * |
|  |  | 5 | 1038.26 | -1954.54 | 11.36/41.16/34.72/11.59/1.18 | 2075.48 | -4020.85 | 1.18/96.18/1.41/0.88/0.35 | * | * | * |
|  | Q | 2 | -2078.55 | 4222.14 | 69.2/30.8 | 2030.63 | -3988.08 | 97.71/2.29 | 2084.9 | -4080.36 | 98.59/1.41 |
|  |  | 3 | -426.64 | 950.85 | 44.69/51.19/4.12 | 2066.43 | -4027.15 | 1.09/96.76/2.15 | 2162.19 | -4202.4 | 1.09/97.94/0.97 |
|  |  | 4 | 626.98 | -1123.86 | 28.86/19.48/50.07/1.59 | 2147.25 | -4156.28 | 1.15/96.26/1.47/1.12 | 2202.44 | -4250.39 | 0.15/1.24/97.71/0.91 |
|  |  | 5 | 1107.57 | -2052.51 | 11.86/34.51/40.95/11.33/1.35 | 2198.37 | -4225.98 | 0.06/3/1.32/94.53/1.09 | 2237.02 | -4287.02 | 0.06/97.38/1.09/1.12/0.35 |
| Uric acid | L | 2 | 3905.09 | -7756.38 | 42.77/57.23 | 12042.25 | -24021.72 | 0.6/99.4 | 12187.18 | -24293.65 | 49.43/50.57 |
|  |  | 3 | 7778.66 | -15476.61 | 19.34/49.38/31.28 | 12261.81 | -24433.94 | 0.34/76.29/23.37 | 12247.95 | -24388.29 | 86.85/13.15/0 |
|  |  | 4 | **9600.61** | **-19093.6** | **7.44/29.57/41.99/20.99** | 12423 | -24729.42 | 95.17/0.34/3.71/0.78 | 12247.95 | -24361.38 | 0/19.82/80.18/0 |
|  |  | 5 | 10610.05 | -21085.59 | 3.06/16.86/32.03/33.58/14.47 | 12476.43 | -24809.37 | 0.33/1.35/2.32/95.49/0.51 | * | * | * |
|  | Q | 2 | 3906.77 | -7741.81 | 42.65/57.35 | 12043.15 | -24005.59 | 0.59/99.41 | 12373.27 | -24647.9 | 99.26/0.74 |
|  |  | 3 | 7780.27 | -15452.93 | 19.36/31.31/49.33 | 12259.41 | -24402.23 | 0.59/96.98/2.43 | 12524.35 | -24914.18 | 0.83/98.46/0.71 |
|  |  | 4 | 9603.46 | -19063.44 | 7.38/41.98/29.57/21.07 | 12406.87 | -24661.3 | 1.11/96.69/0.79/1.41 | 12653.47 | -25136.56 | 0.31/98.36/0.7/0.64 |
|  |  | 5 | 10615.52 | -21051.68 | 3.07/16.93/31.97/33.58/14.46 | 12587.14 | -24985.95 | 0.32/95.74/2.26/0.68/1.01 | 12694.51 | -25182.77 | 0.22/0.69/98.14/0.28/0.68 |
| Albumin | L | 2 | 51969.71 | -103885.49 | 30.87/69.13 | 55585.43 | -111107.93 | 2.85/97.15 | 55711.01 | -111341.11 | 48.37/51.63 |
|  |  | 3 | 53732.52 | -107384.12 | 5.24/47.07/47.68 | 55832.34 | -111574.78 | 0.57/8.48/90.94 | 55972.18 | -111836.47 | 0.69/97.82/1.49 |
|  |  | 4 | 54607.25 | -109106.61 | 1.71/55.11/20.53/22.66 | 55935.33 | -111753.79 | 0.41/4.32/2.47/92.8 | 55918.54 | -111702.21 | 1.28/98.72/0/0 |
|  |  | 5 | 55137.43 | -110140 | 0.34/4.33/26.82/52.69/15.82 | 55973.59 | -111803.34 | 0.41/91.58/3.92/0.36/3.72 | * | * | * |
|  | Q | 2 | 52084.75 | -104097.57 | 30.74/69.26 | 55696.59 | -111312.27 | 2.25/97.75 | 56028.97 | -111959.04 | 1.27/98.73 |
|  |  | 3 | **53873.66** | **-107639.44** | **5.08/47.12/47.8** | 55956.01 | -111795.15 | 0.7/8.78/90.52 | 56180.18 | -112225.5 | 1.08/95.44/3.48 |
|  |  | 4 | 54822.58 | -109501.31 | 1.92/54.57/20.6/22.91 | 56071.57 | -111990.3 | 0.62/6.54/89.31/3.53 | 56261.12 | -112351.42 | 0.73/8.12/90.01/1.13 |
|  |  | 5 | 54982.55 | -109785.29 | 1.82/17.15/36.43/24.56/20.03 | 56186.21 | -112183.62 | 4.95/90.15/1.54/0.4/2.96 | 56420.53 | -112634.27 | 0.21/0.7/87.64/10.45/1 |

C indicates number of classes; TC=total cholesterol; HDL=high-density lipoprotein; LDL=low-density lipoprotein cholesterol; ApoA1=apolipoprotein AI; ApoB=Apolipoprotein B; CRP=C-reactive protein; IgG=immunoglobulin G. The selected model is represented as bold. *Failed to converge; ^a^ Linear models, ^b^ Quadratic models.

**Table S4. Hazard ratios and 95% confidence intervals for the associations of biomarker trajectories with severe infections**

| **Biomarkers** | **Groups** | **N** | **Hospitalization for infectious diseases** | | **Sepsis** | |
| --- | --- | --- | --- | --- | --- | --- |
|  |  |  | **Events/rate, 1000 person-years** | **HRs (95% CI)*** | **Events/rate, 1000 person-years** | **HRs (95% CI)*** |
| Glucose | Class 1: Stable at low levels | 7526 | 2589/38.8 | Ref. | 1160/14.9 | Ref. |
|  | Class 2: Started at high levels and increased | 551 | 188/75.4 | 2.46 (0.62-9.86) | 96/30.9 | 0.59 (0.06-6.13) |
| Fructosamine | Class 1: Stable at low levels | 5994 | 2111/40.8 | Ref. | 952/15.6 | Ref. |
|  | Class 2: Started at high levels and slightly increased | 316 | 112/75.4 | 1.80 (0.24-13.71) | 59/33.6 | 0.42 (0.02-11.01) |
| TC | Class 1: Stable at low levels | 3644 | 1242/38.1 | 0.97 (0.35-2.64) | 590/15.6 | 1.90 (0.59-6.17) |
|  | Class 2: Stable at high levels | 5127 | 1801/41.9 |  | 778/15.4 |  |
| Triglycerides | Class 1: Started at high levels, remained stable, then decreased | 1199 | 440/53.2 | 1.27 (1.14-1.41) | 213/21.4 | 1.26 (1.08-1.46) |
|  | Class 2: Stable at low levels | 7523 | 2589/38.7 | Ref. | 1148/14.7 | Ref. |
| HDL | Class 1: Started at high levels, remained stable, then slightly decreased | 1049 | 403/44.8 | Ref. | 179/17.2 | Ref. |
|  | Class 2: Stable at low levels | 781 | 320/56.4 | 1.26 (1.07-1.49) | 121/17.2 | 0.84 (0.65-1.08) |
| LDL | Class 1: Stable at low levels | 133 | 45/51 | 1.18 (0.84-1.65) | 22/20.8 | 1.42 (0.89-2.28) |
|  | Class 2: Stable at low-middle levels | 447 | 178/56.4 | 1.15 (0.95-1.38) | 70/18.5 | 1.14 (0.85-1.52) |
|  | Class 3: Stable at high-middle levels | 924 | 369/49.9 | Ref. | 160/18.1 | Ref. |
|  | Class 4: Stable at high levels | 465 | 174/43.2 | 0.82 (0.68-1.00) | 73/15.6 | 0.88 (0.66-1.17) |
| LDL/HDL ratio | Class 1: Started at high levels and decreased | 96 | 44/55.3 | 1.04 (0.76-1.43) | 17/17.7 | 0.97 (0.58-1.61) |
|  | Class 2: Stable at low levels | 1733 | 678/48.9 | Ref. | 283/17.2 | Ref. |
| ApoA1 | Class 1: Slight increase followed by a decrease | 991 | 371/49.2 | 1.02 (0.88-1.18) | 154/17.1 | 0.98 (0.78-1.23) |
|  | Class 2: Slight decrease followed by an increase | 995 | 402/49.7 | Ref. | 174/18.2 | Ref. |
| ApoB | Class 1: Started at high levels and decreased | 109 | 50/62.9 | 1.25 (0.93-1.68) | 24/25.1 | 1.40 (0.91-2.17) |
|  | Class 2: Stable at low levels | 1798 | 692/48.1 | Ref. | 290/17 | Ref. |
| ApoB/ApoA1 ratio | Class 1: Started at high levels and decreased | 182 | 79/56.8 | 1.12 (0.88-1.42) | 29/16.4 | 0.80 (0.53-1.20) |
|  | Class 2: Stable at low levels | 1612 | 618/49.4 | Ref. | 263/17.8 | Ref. |
| Hemoglobin | Class 1: Started at low levels, remained stable, then decreased | 244 | 73/41.2 | 0.87 (0.67-1.12) | 37/17.7 | 1.33 (0.92-1.91) |
|  | Class 2: Stable at low-middle levels | 1619 | 630/53.3 | Ref. | 252/17.4 | Ref. |
|  | Class 3: Stable at high-middle levels | 967 | 325/44.3 | 0.87 (0.75-1.01) | 125/14.2 | 1.03 (0.81-1.30) |
|  | Class 4: Stable at high levels | 790 | 265/49.2 | 0.93 (0.80-1.09) | 109/17 | 0.80 (0.63-1.02) |
| Globulin | Class 1: Stable at low levels | 1203 | 479/46.5 | Ref. | 183/14.7 | Ref. |
|  | Class 2: Started at high levels and slightly increased | 589 | 199/52.8 | 1.09 (0.91-1.29) | 86/19.3 | 1.28 (0.98-1.67) |
| Haptoglobin | Class 1: Stable at low levels | 816 | 297/34.5 | 1.00 (0.87-1.14) | 136/13.5 | 1.07 (0.88-1.31) |
|  | Class 2: Stable at high levels | 1379 | 418/51.1 | 1.38 (1.23-1.55) | 188/19.2 | 1.42 (1.19-1.70) |
|  | Class 3: Stable at intermediate levels | 3015 | 1029/36.5 | Ref. | 440/13.4 | Ref. |
| CRP | Class 1: Started at low levels, remained stable, then increased rapidly | 489 | 178/54.9 | 1.24 (1.06-1.46) | 90/23.8 | 1.43 (1.14-1.79) |
|  | Class 2: Started at relatively higher levels but remained stable | 3590 | 1311/43.1 | Ref. | 579/16.1 | Ref. |
| IgG | Class 1: Stable at low levels | 504 | 178/47.3 | Ref. | 69/15.4 | Ref. |
|  | Class 2: Started at low levels and increased rapidly | 30 | 9/85.2 | 1.78 (0.82-3.85) | 7/49.6 | 3.39 (1.43-8.07) |
| Leukocyte | Class 1: Stable at intermediate levels | 1614 | 590/58.4 | Ref. | 226/18.1 | Ref. |
|  | Class 2: Stable at low levels | 1547 | 565/43.3 | 0.40 (0.08-1.98) | 227/14.5 | 0.19 (0.03-1.36) |
|  | Class 3: Started at high levels and increased | 238 | 75/61.6 | 0.01 (0-0.45) | 29/19.5 | 0 (0-6.23) |
| Uric acid | Class 1: Stable at low levels | 584 | 197/29.2 | 0.83 (0.71-0.99) | 81/10.4 | 0.86 (0.67-1.12) |
|  | Class 2: Stable at low-middle levels | 2320 | 784/34 | 0.89 (0.80-0.98) | 307/11.4 | 0.85 (0.73-0.99) |
|  | Class 3: Stable at high-middle levels | 3294 | 1124/43.3 | Ref. | 516/16.8 | Ref. |
|  | Class 4: Stable at high levels | 1647 | 610/57.3 | 1.19 (1.07-1.32) | 319/25.5 | 1.36 (1.18-1.58) |
| Albumin | Class 1: Started at low levels, remained stable, then decreased | 408 | 112/56.9 | 1.22 (1.00-1.50) | 63/27.4 | 1.65 (1.26-2.16) |
|  | Class 2: Stable at intermediate levels | 3783 | 1267/43.6 | 1.01 (0.93-1.10) | 560/16.4 | 1.07 (0.95-1.20) |
|  | Class 3: Stable at high levels | 3837 | 1380/39.5 | Ref. | 631/15.4 | Ref. |

HR=hazard ratio; CI=confidence intervals; TC=total cholesterol; HDL=high-density lipoprotein; LDL=low-density lipoprotein; ApoA1=apolipoprotein A1; ApoB=apolipoprotein B; CRP=C-reactive protein; IgG=immunoglobulin G.

*Adjusted for sex, calendar year of cancer diagnosis, age at cancer diagnosis, country of birth, education, income, and employment status at the first blood sampling.

**Table S5. Hazard ratios and 95% confidence intervals for the associations of biomarker trajectories with severe infections after additionally adjusting for comorbidities**

| **Biomarkers** | **Groups** | **Hospitalization for infectious diseases** | **Sepsis** |
| --- | --- | --- | --- |
| Glucose | Class 1: Stable at low levels | Ref. | Ref. |
|  | Class 2: Started at high levels and increased | 2.10 (0.52-8.40) | 0.53 (0.05-5.44) |
| Fructosamine | Class 1: Stable at low levels | Ref. | Ref. |
|  | Class 2: Started at high levels and slightly increased | 1.51 (0.20-11.41) | 0.39 (0.02-10.18) |
| TC | Class 1: Stable at low levels | 0.94 (0.34-2.56) | 1.84 (0.57-5.97) |
|  | Class 2: Stable at high levels | Ref. | Ref. |
| Triglycerides | Class 1: Started at high levels, remained stable, then decreased | 1.20 (1.08-1.33) | 1.19 (1.02-1.39) |
|  | Class 2: Stable at low levels | Ref. | Ref. |
| HDL | Class 1: Started at high levels, remained stable, then slightly decreased | Ref. | Ref. |
|  | Class 2: Stable at low levels | 1.20 (1.01-1.42) | 0.78 (0.60-1.01) |
| LDL | Class 1: Stable at low levels | 1.15 (0.82-1.60) | 1.40 (0.87-2.24) |
|  | Class 2: Stable at low-middle levels | 1.14 (0.94-1.37) | 1.11 (0.83-1.48) |
|  | Class 3: Stable at high-middle levels | Ref. | Ref. |
|  | Class 4: Stable at high levels | 0.85 (0.70-1.03) | 0.90 (0.67-1.20) |
| LDL/HDL ratio | Class 1: Started at high levels and decreased | 1.01 (0.74-1.38) | 0.93 (0.56-1.54) |
|  | Class 2: Stable at low levels | Ref. | Ref. |
| ApoA1 | Class 1: Slight increase followed by a decrease | 1.02 (0.88-1.19) | 0.98 (0.79-1.23) |
|  | Class 2: Slight decrease followed by an increase | Ref. | Ref. |
| ApoB | Class 1: Started at high levels and decreased | 1.25 (0.93-1.69) | 1.38 (0.89-2.14) |
|  | Class 2: Stable at low levels | Ref. | Ref. |
| ApoB/ApoA1 ratio | Class 1: Started at high levels and decreased | 1.10 (0.87-1.40) | 0.76 (0.51-1.15) |
|  | Class 2: Stable at low levels | Ref. | Ref. |
| Hemoglobin | Class 1: Started at low levels, remained stable, then decreased | 0.87 (0.67-1.13) | 1.32 (0.92-1.89) |
|  | Class 2: Stable at low-middle levels | Ref. | Ref. |
|  | Class 3: Stable at high-middle levels | 0.87 (0.75-1.00) | 1.02 (0.81-1.29) |
|  | Class 4: Stable at high levels | 0.93 (0.80-1.09) | 0.79 (0.62-1.01) |
| Globulin | Class 1: Stable at low levels | Ref. | Ref. |
|  | Class 2: Started at high levels and slightly increased | 1.02 (0.86-1.22) | 1.22 (0.93-1.60) |
| Haptoglobin | Class 1: Stable at low levels | 1.01 (0.88-1.15) | 1.08 (0.89-1.32) |
|  | Class 2: Stable at high levels | 1.32 (1.17-1.49) | 1.39 (1.16-1.66) |
|  | Class 3: Stable at intermediate levels | Ref. | Ref. |
| CRP | Class 1: Started at low levels, remained stable, then increased rapidly | 1.23 (1.04-1.44) | 1.41 (1.12-1.77) |
|  | Class 2: Started at relatively higher levels but remained stable | Ref. | Ref. |
| IgG | Class 1: Stable at low levels | Ref. | Ref. |
|  | Class 2: Started at low levels and increased rapidly | 1.78 (0.81-3.91) | 3.25 (1.33-7.96) |
| Leukocyte | Class 1: Stable at intermediate levels | Ref. | Ref. |
|  | Class 2: Stable at low levels | 0.41 (0.08-2.03) | 0.20 (0.03-1.42) |
|  | Class 3: Started at high levels and increased | 0 (0-0.43) | 0 (0-7.07) |
| Uric acid | Class 1: Stable at low levels | 0.84 (0.72-1.00) | 0.87 (0.67-1.12) |
|  | Class 2: Stable at low-middle levels | 0.88 (0.80-0.98) | 0.84 (0.72-0.99) |
|  | Class 3: Stable at high-middle levels | Ref. | Ref. |
|  | Class 4: Stable at high levels | 1.16 (1.04-1.29) | 1.32 (1.14-1.53) |
| Albumin | Class 1: Started at low levels, remained stable, then decreased | 1.16 (0.94-1.42) | 1.60 (1.22-2.10) |
|  | Class 2: Stable at intermediate levels | 1.02 (0.94-1.10) | 1.07 (0.95-1.20) |
|  | Class 3: Stable at high levels | Ref. | Ref. |

TC=total cholesterol; HDL=high-density lipoprotein; LDL=low-density lipoprotein; ApoA1=apolipoprotein A1; ApoB=apolipoprotein B; CRP=C-reactive protein; IgG=immunoglobulin G.

Adjusted for sex, calendar year of cancer diagnosis, age at cancer diagnosis, country of birth, education, income, and employment status at the first blood sampling, as well as a history of severe infections within 10 years before cancer diagnosis, psychiatric disorders and diabetes before cancer diagnosis.

**Table S6. Hazard ratios and 95% confidence intervals for the associations between biomarker trajectories and hospitalization for infectious diseases defined by both primary and secondary diagnoses from the Patient Register**

| **Biomarkers** | **Groups** | **Event/rate, per 1000 person-years** | **HRs (95%CI)** |
| --- | --- | --- | --- |
| Glucose | Class 1: Stable at low levels | 3978/67.6 | Ref. |
|  | Class 2: Started at high levels and increased | 281/128.5 | 0.65 (0.24-1.77) |
| Fructosamine | Class 1: Stable at low levels | 3231/71.3 | Ref. |
|  | Class 2: Started at high levels and slightly increased | 166/128.6 | 0.36 (0.08-1.73) |
| TC | Class 1: Stable at low levels | 1868/64.3 | 0.85 (0.50-1.45) |
|  | Class 2: Stable at high levels | 2784/74 | Ref. |
| Triglycerides | Class 1: Started at high levels, remained stable, then decreased | 665/93.3 | 1.22 (1.12-1.33) |
|  | Class 2: Stable at low levels | 3963/66.9 | Ref. |
| HDL | Class 1: Started at high levels, remained stable, then slightly decreased | 590/74.8 | Ref. |
|  | Class 2: Stable at low levels | 463/94.9 | 1.22 (1.07-1.40) |
| LDL | Class 1: Stable at low levels | 66/82.2 | 1.07 (0.81-1.40) |
|  | Class 2: Stable at low-middle levels | 251/89.3 | 1.04 (0.89-1.22) |
|  | Class 3: Stable at high-middle levels | 541/85.8 | Ref. |
|  | Class 4: Stable at high levels | 267/74.9 | 0.86 (0.74-1.01) |
| LDL/HDL ratio | Class 1: Started at high levels and decreased | 64/91.7 | 1.00 (0.77-1.30) |
|  | Class 2: Stable at low levels | 988/81.9 | Ref. |
| ApoA1 | Class 1: Slight increase followed by a decrease | 565/84.8 | 1.05 (0.93-1.19) |
|  | Class 2: Slight decrease followed by an increase | 570/81.7 | Ref. |
| ApoB | Class 1: Started at high levels and decreased | 70/111.3 | 1.23 (0.95-1.58) |
|  | Class 2: Stable at low levels | 1032/81.8 | Ref. |
| ApoB/ApoA1 ratio | Class 1: Started at high levels and decreased | 112/95.9 | 1.05 (0.86-1.28) |
|  | Class 2: Stable at low levels | 921/84.3 | Ref. |
| Hemoglobin | Class 1: Started at low levels, remained stable, then decreased | 134/92.2 | 1.17 (0.96-1.43) |
|  | Class 2: Stable at low-middle levels | 930/90.1 | Ref. |
|  | Class 3: Stable at high-middle levels | 521/82.6 | 1.02 (0.91-1.15) |
|  | Class 4: Stable at high levels | 416/87.6 | 0.97 (0.86-1.10) |
| Globulin | Class 1: Stable at low levels | 683/74.5 | Ref. |
|  | Class 2: Started at high levels and slightly increased | 306/93.4 | 1.13 (0.98-1.30) |
| Haptoglobin | Class 1: Stable at low levels | 426/54.9 | 0.94 (0.84-1.05) |
|  | Class 2: Stable at high levels | 670/94.8 | 1.40 (1.27-1.54) |
|  | Class 3: Stable at intermediate levels | 1578/62.9 | Ref. |
| CRP | Class 1: Started at low levels, remained stable, then increased rapidly | 265/95.6 | 1.17 (1.03-1.34) |
|  | Class 2: Started at relatively higher levels but remained stable | 1990/74.9 | Ref. |
| IgG | Class 1: Stable at low levels | 268/81 | Ref. |
|  | Class 2: Started at low levels and increased rapidly | 17/247.9 | 2.39 (1.38-4.15) |
| Leukocyte | Class 1: Stable at intermediate levels | 919/106.8 | Ref. |
|  | Class 2: Stable at low levels | 862/75.4 | 0.62 (0.29-1.33) |
|  | Class 3: Started at high levels and increased | 122/122.7 | 0.35 (0.06-1.89) |
| Uric acid | Class 1: Stable at low levels | 290/48 | 0.80 (0.69-0.91) |
|  | Class 2: Stable at low-middle levels | 1151/55.2 | 0.83 (0.76-0.90) |
|  | Class 3: Stable at high-middle levels | 1774/78.5 | Ref. |
|  | Class 4: Stable at high levels | 937/103 | 1.14 (1.05-1.24) |
| Albumin | Class 1: Started at low levels, remained stable, then decreased | 198/117.5 | 1.30 (1.11-1.52) |
|  | Class 2: Stable at intermediate levels | 2002/78.3 | 1.04 (0.98-1.11) |
|  | Class 3: Stable at high levels | 2066/67.7 | Ref. |

HR=hazard ratio; CI=confidence intervals; TC=total cholesterol; HDL=high-density lipoprotein; LDL=low-density lipoprotein; ApoA1=apolipoprotein A1; ApoB=apolipoprotein B; CRP=C-reactive protein; IgG=immunoglobulin G.

Adjusted for sex, calendar year of cancer diagnosis, age at cancer diagnosis, country of birth, education, income, and employment status at the first blood sampling.

**Table S7. Hazard ratios and 95% confidence intervals for the associations between biomarker trajectories and hospitalization for infectious diseases stratified by sex**

| **Biomarkers** | **Reference** | **Groups** | **Men** | **Women** |
| --- | --- | --- | --- | --- |
| Glucose | Class 1: Stable at low levels | Class 2: Started at high levels and increased | 1.44 (0.20-10.13) | 4.90 (0.68-35.32) |
| Fructosamine | Class 1: Stable at low levels | Class 2: Started at high levels and slightly increased | 0.77 (0.04-13.83) | 5.06 (0.31-82.11) |
| TC | Class 2: Stable at high levels | Class 1: Stable at low levels | 0.72 (0.18-2.86) | 1.42 (0.33-6.14) |
| Triglycerides | Class 2: Stable at low levels | Class 1: Started at high levels, remained stable, then decreased | 1.15 (1.01-1.32) | 1.52 (1.28-1.81) |
| HDL | Class 1: Started at high levels, remained stable, then slightly decreased | Class 2: Stable at low levels | 1.05 (0.85-1.30) | 1.63 (1.28-2.09) |
| LDL | Class 3: Started at high-middle levels | Class 1: Started at low levels | 1.22 (0.74-2.01) | 1.15 (0.73-1.83) |
|  |  | Class 2: Started at low-middle levels | 1.19 (0.92-1.54) | 1.12 (0.85-1.49) |
|  |  | Class 4: Started at high levels | 0.81 (0.63-1.05) | 0.84 (0.64-1.11) |
| LDL/HDL ratio | Class 2: Stable at low levels | Class 1: Started at high levels and decreased | 1.31 (0.83-2.05) | 0.92 (0.59-1.45) |
| ApoA1 | Class 2: Slight decrease followed by an increase | Class 1: Slight increase followed by a decrease | 0.99 (0.81-1.21) | 1.05 (0.84-1.30) |
| ApoB | Class 2: Stable at low levels | Class 1: Started at high levels and decreased | 1.39 (0.90-2.15) | 1.14 (0.75-1.72) |
| ApoB/ApoA1 ratio | Class 2: Stable at low levels | Class 1: Started at high levels and decreased | 1.07 (0.75-1.52) | 1.13 (0.81-1.59) |
| Hemoglobin | Class 2: Stable at low-middle levels | Class 1: Started at low levels, remained stable, then decreased | 1.01 (0.56-1.80) | 0.83 (0.62-1.11) |
|  |  | Class 3: Stable at high-middle levels | 0.96 (0.69-1.32) | 0.86 (0.73-1.02) |
|  |  | Class 4: Stable at high levels | 0.90 (0.75-1.08) | 1.06 (0.76-1.47) |
| Globulin | Class 1: Stable at low levels | Class 2: Started at high levels and slightly increased | 1.20 (0.94-1.52) | 0.98 (0.76-1.25) |
| Haptoglobin | Class 3: Stable at intermediate levels | Class 1: Stable at low levels | 0.94 (0.78-1.13) | 1.05 (0.87-1.27) |
|  |  | Class 2: Stable at high levels | 1.20 (1.02-1.41) | 1.60 (1.35-1.90) |
| CRP | Class 2: Started at relatively higher levels but remained stable | Class 1: Started at low levels, remained stable, then increased rapidly | 1.31 (1.05-1.63) | 1.18 (0.93-1.49) |
| IgG | Class 1: Stable at low levels | Class 2: Started at low levels and increased rapidly | 2.94 (1.02-8.52) | 1.61 (0.49-5.25) |
| Leukocyte | Class 1: Stable at intermediate levels | Class 2: Stable at low levels | 1.10 (0.10-11.65) | 0.17 (0.02-1.46) |
|  |  | Class 3: Started at high levels and increased | 0 (0-23.83) | 0.01 (0-0.94) |
| Uric acid | Class 3: Stable at high-middle levels | Class 1: Stable at low levels | 1.32 (0.79-2.21) | 0.79 (0.66-0.95) |
|  |  | Class 2: Stable at low-middle levels | 0.99 (0.84-1.16) | 0.84 (0.74-0.96) |
|  |  | Class 4: Stable at high levels | 1.16 (1.03-1.31) | 1.33 (1.08-1.63) |
| Albumin | Class 3: Stable at high levels | Class 1: Started at low levels, remained stable, then decreased | 1.23 (0.91-1.67) | 1.19 (0.90-1.57) |
|  |  | Class 2: Stable at intermediate levels | 1.06 (0.94-1.19) | 0.97 (0.87-1.09) |

TC=total cholesterol; HDL=high-density lipoprotein; LDL=low-density lipoprotein; ApoA1=apolipoprotein A1; ApoB=apolipoprotein B; CRP=C-reactive protein; IgG=immunoglobulin G.

Adjusted for sex, calendar year of cancer diagnosis, age at cancer diagnosis, country of birth, education, income, and employment status at the first blood sampling.

**Table S8. Hazard ratios and 95% confidence intervals for the associations between biomarker trajectories and hospitalization for infectious diseases stratified by age at cancer diagnosis**

| **Biomarkers** | **Reference** | **Groups** | **≤ 60 years** | **> 60 years** |
| --- | --- | --- | --- | --- |
| Glucose | Class 1: Stable at low levels | Class 2: Started at high levels and increased | 4.52 (0.50-40.9) | 2.29 (0.38-13.9) |
| Fructosamine | Class 1: Stable at low levels | Class 2: Started at high levels and slightly increased | 3.32 (0.16-67.69) | 1.12 (0.07-18.32) |
| TC | Class 2: Stable at high levels | Class 1: Stable at low levels | 0.60 (0.14-2.63) | 1.12 (0.30-4.28) |
| Triglycerides | Class 2: Stable at low levels | Class 1: Started at high levels, remained stable, then decreased | 1.24 (1.03-1.50) | 1.31 (1.15-1.48) |
| HDL | Class 1: Started at high levels, remained stable, then slightly decreased | Class 2: Stable at low levels | 1.21 (0.86-1.71) | 1.31 (1.09-1.59) |
| LDL | Class 3: Started at high-middle levels | Class 1: Started at low levels | 1.18 (0.67-2.07) | 1.20 (0.78-1.84) |
|  |  | Class 2: Started at low-middle levels | 1.52 (1.06-2.18) | 1.02 (0.81-1.27) |
|  |  | Class 4: Started at high levels | 1.06 (0.73-1.54) | 0.75 (0.61-0.94) |
| LDL/HDL ratio | Class 2: Stable at low levels | Class 1: Started at high levels and decreased | 1.13 (0.55-2.32) | 1.05 (0.74-1.49) |
| ApoA1 | Class 2: Slight decrease followed by an increase | Class 1: Slight increase followed by a decrease | 0.96 (0.72-1.27) | 1.01 (0.85-1.20) |
| ApoB | Class 2: Stable at low levels | Class 1: Started at high levels and decreased | 1.49 (0.77-2.86) | 1.19 (0.85-1.67) |
| ApoB/ApoA1 ratio | Class 2: Stable at low levels | Class 1: Started at high levels and decreased | 1.35 (0.83-2.21) | 1.07 (0.82-1.42) |
| Hemoglobin | Class 2: Stable at low-middle levels | Class 1: Started at low levels, remained stable, then decreased | 0.65 (0.44-0.98) | 1.11 (0.79-1.57) |
|  |  | Class 3: Stable at high-middle levels | 0.71 (0.54-0.93) | 0.94 (0.79-1.11) |
|  |  | Class 4: Stable at high levels | 0.92 (0.66-1.29) | 0.95 (0.79-1.13) |
| Globulin | Class 1: Stable at low levels | Class 2: Started at high levels and slightly increased | 1.16 (0.83-1.63) | 1.06 (0.87-1.30) |
| Haptoglobin | Class 3: Stable at intermediate levels | Class 1: Stable at low levels | 0.85 (0.68-1.06) | 1.11 (0.94-1.31) |
|  |  | Class 2: Stable at high levels | 1.39 (1.13-1.70) | 1.40 (1.21-1.62) |
| CRP | Class 2: Started at relatively higher levels but remained stable | Class 1: Started at low levels, remained stable, then increased rapidly | 1.25 (0.92-1.69) | 1.24 (1.02-1.50) |
| IgG | Class 1: Stable at low levels | Class 2: Started at low levels and increased rapidly | 1.31 (0.15-11.04) | 1.84 (0.78-4.35) |
| Leukocyte | Class 1: Stable at intermediate levels | Class 2: Stable at low levels | 0.26 (0.01-4.58) | 0.41 (0.06-2.82) |
|  |  | Class 3: Started at high levels and increased | 0.09 (0-19.22) | 0 (0-0.25) |
| Uric acid | Class 3: Stable at high-middle levels | Class 1: Stable at low levels | 0.69 (0.54-0.88) | 0.96 (0.76-1.21) |
|  |  | Class 2: Stable at low-middle levels | 0.78 (0.66-0.93) | 0.94 (0.83-1.06) |
|  |  | Class 4: Stable at high levels | 1.07 (0.87-1.32) | 1.24 (1.10-1.40) |
| Albumin | Class 3: Stable at high levels | Class 1: Started at low levels, remained stable, then decreased | 1.46 (0.96-2.23) | 1.16 (0.92-1.46) |
|  |  | Class 2: Stable at intermediate levels | 1.04 (0.90-1.20) | 1.01 (0.91-1.11) |

TC=total cholesterol; HDL=high-density lipoprotein; LDL=low-density lipoprotein; ApoA1=apolipoprotein A1; ApoB=apolipoprotein B; CRP=C-reactive protein; IgG=immunoglobulin G.

Adjusted for sex, calendar year of cancer diagnosis, age at cancer diagnosis, country of birth, education, income, and employment status at the first blood sampling.

**Table S9. Hazard ratios and 95% confidence intervals for the associations between biomarker trajectories and hospitalization for infectious diseases stratified by a history of severe infections during the 10 years before cancer diagnosis**

| **Biomarkers** | **Reference** | **Groups** | **History of severe infections** | |
| --- | --- | --- | --- | --- |
|  |  |  | **No** | **Yes** |
| Glucose | Class 1: Stable at low levels | Class 2: Started at high levels and increased | 0.99 (0.15-6.59) | 5.81 (0.67-50.61) |
| Fructosamine | Class 1: Stable at low levels | Class 2: Started at high levels and slightly increased | 1.14 (0.10-13.65) | 3.34 (0.14-79.56) |
| TC | Class 2: Stable at high levels | Class 1: Stable at low levels | 1.21 (0.40-3.66) | 0.21 (0.02-2.57) |
| Triglycerides | Class 2: Stable at low levels | Class 1: Started at high levels, remained stable, then decreased | 1.26 (1.12-1.41) | 1.22 (0.93-1.60) |
| HDL | Class 1: Started at high levels, remained stable, then slightly decreased | Class 2: Stable at low levels | 1.24 (1.03-1.48) | 1.16 (0.74-1.82) |
| LDL | Class 3: Started at high-middle levels | Class 1: Started at low levels | 1.18 (0.82-1.71) | 1.01 (0.45-2.27) |
|  |  | Class 2: Started at low-middle levels | 1.14 (0.93-1.39) | 1.16 (0.70-1.91) |
|  |  | Class 4: Started at high levels | 0.80 (0.65-0.97) | 1.19 (0.71-2.00) |
| LDL/HDL ratio | Class 2: Stable at low levels | Class 1: Started at high levels and decreased | 0.96 (0.67-1.37) | 1.53 (0.78-3.01) |
| ApoA1 | Class 2: Slight decrease followed by an increase | Class 1: Slight increase followed by a decrease | 1.00 (0.85-1.17) | 1.15 (0.75-1.74) |
| ApoB | Class 2: Stable at low levels | Class 1: Started at high levels and decreased | 1.24 (0.89-1.71) | 1.57 (0.71-3.47) |
| ApoB/ApoA1 ratio | Class 2: Stable at low levels | Class 1: Started at high levels and decreased | 1.04 (0.80-1.36) | 1.54 (0.88-2.69) |
| Hemoglobin | Class 2: Stable at low-middle levels | Class 1: Started at low levels, remained stable, then decreased | 0.87 (0.65-1.16) | 0.79 (0.41-1.51) |
|  |  | Class 3: Stable at high-middle levels | 0.83 (0.70-0.97) | 1.10 (0.77-1.56) |
|  |  | Class 4: Stable at high levels | 0.92 (0.77-1.09) | 1.02 (0.68-1.52) |
| Globulin | Class 1: Stable at low levels | Class 2: Started at high levels and slightly increased | 1.05 (0.87-1.28) | 1.12 (0.73-1.73) |
| Haptoglobin | Class 3: Stable at intermediate levels | Class 1: Stable at low levels | 0.97 (0.84-1.12) | 1.27 (0.85-1.88) |
|  |  | Class 2: Stable at high levels | 1.37 (1.20-1.55) | 1.32 (0.95-1.83) |
| CRP | Class 2: Started at relatively higher levels but remained stable | Class 1: Started at low levels, remained stable, then increased rapidly | 1.19 (0.99-1.42) | 1.57 (1.08-2.28) |
| IgG | Class 1: Stable at low levels | Class 2: Started at low levels and increased rapidly | 1.93 (0.78-4.80) | 2.08 (0.39-11.23) |
| Leukocyte | Class 1: Stable at intermediate levels | Class 2: Stable at low levels | 0.51 (0.10-2.71) | 0.05 (0-5.96) |
|  |  | Class 3: Started at high levels and increased | 0.01 (0-1.35) | 0 (0-28.93) |
| Uric acid | Class 3: Stable at high-middle levels | Class 1: Stable at low levels | 0.80 (0.67-0.95) | 1.41 (0.89-2.23) |
|  |  | Class 2: Stable at low-middle levels | 0.87 (0.78-0.97) | 0.99 (0.74-1.33) |
|  |  | Class 4: Stable at high levels | 1.19 (1.06-1.33) | 1.10 (0.83-1.46) |
| Albumin | Class 3: Stable at high levels | Class 1: Started at low levels, remained stable, then decreased | 1.19 (0.95-1.49) | 1.25 (0.78-2.00) |
|  |  | Class 2: Stable at intermediate levels | 1.00 (0.92-1.09) | 1.03 (0.82-1.28) |

TC=total cholesterol; HDL=high-density lipoprotein; LDL=low-density lipoprotein; ApoA1=apolipoprotein A1; ApoB=apolipoprotein B; CRP=C-reactive protein; IgG=immunoglobulin G.

*Only a few number of outcome events were observed.

Adjusted for sex, calendar year of cancer diagnosis, age at cancer diagnosis, country of birth, education, income, and employment status at the first blood sampling.

**Table S10. Hazard ratios and 95% confidence intervals for the associations between biomarker trajectories and hospitalization for infectious diseases among patients with breast and reproductive system cancers, digestive system cancers, and hematological malignancies, respectively***

| **Biomarkers** | **Reference** | **Groups** | **Breast and reproductive system cancers** | **Digestive system cancers** | **Hematological malignancies** |
| --- | --- | --- | --- | --- | --- |
| Triglycerides | Class 2: Stable at low levels | Class 1: Started at high levels, remained stable, then decreased | 1.21 (1.03-1.43) | 1.23 (0.93-1.62) | 1.44 (1.03-1.99) |
| HDL | Class 1: Started at high levels, remained stable, then slightly decreased | Class 2: Stable at low levels | 1.17 (0.92-1.48) | 1.11 (0.71-1.73) | 1.15 (0.66-2.00) |
| Haptoglobin | Class 3: Stable at intermediate levels | Class 1: Stable at low levels | 0.97 (0.79-1.19) | 1.24 (0.82-1.86) | 1.03 (0.70-1.50) |
|  |  | Class 2: Stable at high levels | 1.44 (1.20-1.74) | 1.28 (0.93-1.77) | 1.10 (0.74-1.65) |
| CRP | Class 2: Started at relatively higher levels but remained stable | Class 1: Started at low levels, remained stable, then increased rapidly | 1.35 (1.05-1.74) | 1.28 (0.86-1.91) | 1.19 (0.70-2.03) |
| Uric acid | Class 3: Stable at high-middle levels | Class 1: Stable at low levels | 0.77 (0.61-0.98) | 0.88 (0.54-1.45) | 0.98 (0.59-1.62) |
|  |  | Class 2: Stable at low-middle levels | 0.85 (0.73-0.98) | 1.19 (0.89-1.59) | 0.74 (0.53-1.03) |
|  |  | Class 4: Stable at high levels | 1.18 (1.00-1.39) | 1.36 (1.03-1.81) | 1.08 (0.78-1.50) |
| Albumin | Class 3: Stable at high levels | Class 1: Started at low levels, remained stable, then decreased | 1.12 (0.80-1.56) | 0.92 (0.53-1.57) | 0.99 (0.59-1.66) |
|  |  | Class 2: Stable at intermediate levels | 0.97 (0.86-1.10) | 1.09 (0.87-1.37) | 1.03 (0.79-1.33) |

*Due to the small number of patients classified by cancer types, we only focused on triglycerides, high-density lipoprotein, haptoglobin, C-reactive protein, uric acid, and albumin.

TC=total cholesterol; HDL=high-density lipoprotein; LDL=low-density lipoprotein; ApoA1=apolipoprotein A1; ApoB=apolipoprotein B; CRP=C-reactive protein; IgG=immunoglobulin G.

Adjusted for sex, calendar year of cancer diagnosis, age at cancer diagnosis, country of birth, education, income, and employment status at the first blood sampling.

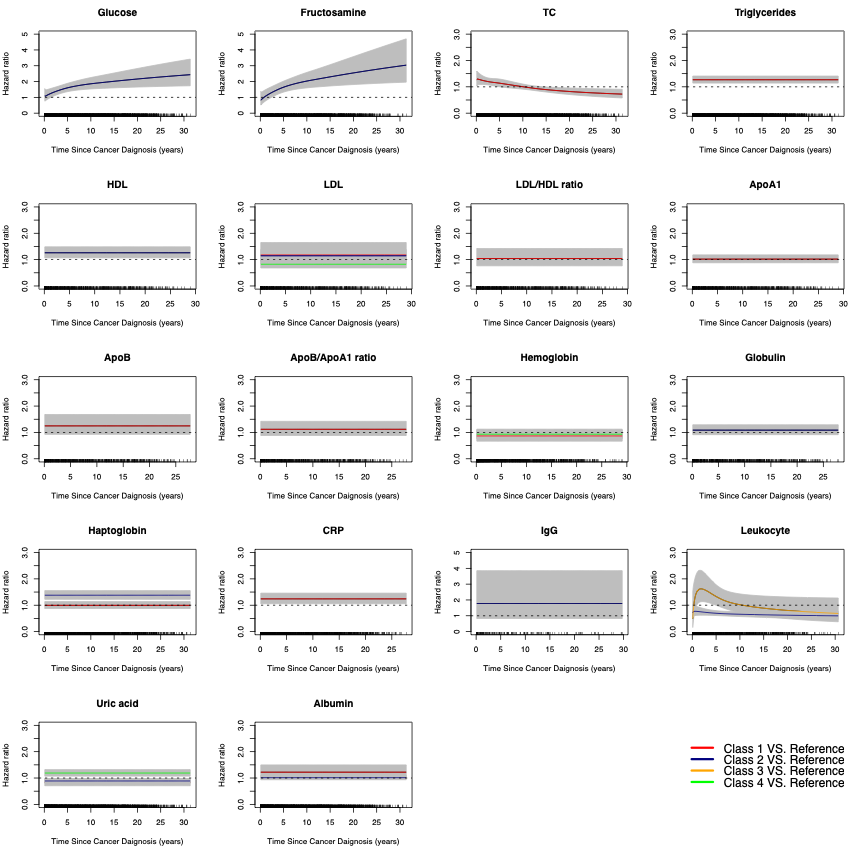

**Figure S1. Crude hazard ratios and 95% confidence intervals of hospitalization for infectious diseases in relation to biomarker trajectories**

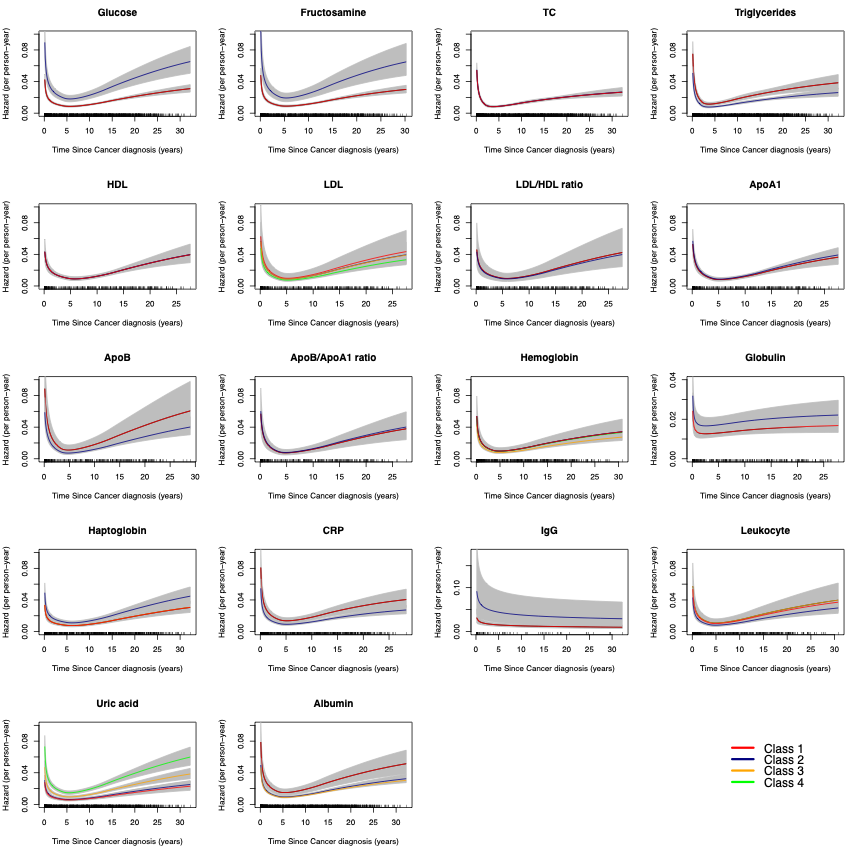

**Figure S2. Hazards of sepsis diagnosis after a cancer diagnosis in relation to biomarker trajectories**
